# Validity of a Common Measure of Intimate Partner Violence Perpetration: Impact on Study Inference in Trials in Low- and Middle-Income Countries

**DOI:** 10.1101/2024.01.28.24301897

**Authors:** Cari Jo Clark, Irina Bergenfeld, Abbie Shervinskie, Erin R. Johnson, Yuk Fai Cheong, Nadine J Kaslow, Kathryn M Yount

**Author notes:** **Corresponding author**: Cari Jo Clark, 1518 Clifton Road, NE, Atlanta, Georgia, 30322.

## Abstract

**Background:** In lower-and middle-income countries (LMICs), studies of interventions to reduce intimate partner violence (IPV) perpetration are expanding, yet measurement equivalence of the IPV perpetration construct that is the primary outcome in these investigations has not been established. We assessed the measurement equivalence of physical and sexual IPV perpetration item sets used in recent trials in LMICs and tested the impact of non-invariance on trial inference.

**Methods:** With data from three intervention trials among men (sample size 505-1537 across studies) completed in 2019, we calculated tetrachoric correlations among items and used multiple-group confirmatory factor analysis to assess invariance across arms and over time. We also assessed treatment effects adjusting for covariate imbalance and using inverse probability to treatment weights to assess concordance of invariant measures with published results, where warranted.

**Findings:** The average correlation among items measuring IPV perpetration was high and increased by 0.03 to 0.15 for physical IPV and 0.07 to 0.17 for sexual IPV over time with several items in two studies showing correlations ≥ 0.85 at endline. Increases in the degree of correlation for physical IPV were concentrated in the treatment arm in two of the studies. The increase in correlation in sexual IPV differed by arm across studies. Across all studies, a correlated two-factor solution was the best fitting model according to the EFAs and CFAs. One study demonstrated measurement invariance across arms and over time. In two of the studies, longitudinal measurement non-invariance was detected in the intervention arms. In post hoc testing, one study attained invariance with a one-factor model and study inference was concordant with published findings. The other study did not attain even partial invariance.

**Conclusion:** Common measures of physical and sexual IPV perpetration cannot be used validly for comparisons across treatment versus control groups over time without further refinement. The study highlights the need for an expanded item set, content validity assessments, further measurement invariance testing, and then consistent use of the item sets in future intervention trials to ensure valid inferences regarding the effectiveness of IPV perpetration prevention interventions within and across trials.

## INTRODUCTION

Intimate partner violence (IPV) remains a critical public health concern due to its high morbidity and mortality. As a result, interest is growing to develop and evaluate prevention interventions aimed at reducing men’s perpetration of IPV globally. While there has been growth in assessing the effectiveness of these interventions, intervention effectiveness cannot be optimally examined until there is accurate measurement of IPV perpetration. Making valid comparisons of the mean or prevalence of IPV perpetration across study arms and intervention trials requires “cross-group” measurement invariance meaning that respondents perceive and respond similarly to the scale items regardless of the “group” to which they belong. Moreover, making valid comparisons of the mean or prevalence of perpetration over time requires “cross-time” invariance meaning that respondents perceive and respond similarly to the scale items over time. Cross-group, cross-trial, and cross-time measurement invariance ensures that comparisons of the effects of primary intervention interventions on IPV perpetration can be validly made.

Despite growing research in the field of violence prevention, comprehensive assessment of the measurement invariance of commonly used scales of IPV perpetration has been absent. Existing cross-sectional studies have identified non invariance (I.e., inadequate model fit with or without modifications) (Putnick and Bornstein 2016) or partial invariance (adequate model fit only after modification) (Putnick and Bornstein 2016) of perpetration scales across genders (O’Hara, Perkins et al. 2018, Shorey, Allan et al. 2019, Wareham, Wagers et al. 2022), language of administration (Connelly, Newton et al. 2005), and race/ethnicity (Shorey, Allan et al. 2019). Collectively, the existing research suggests a potential lack of comparability in the construct of IPV perpetration, raising concerns regarding the validity of comparisons made across intervention and control groups, over time, and across different populations. To the authors’ knowledge, no study has examined measurement invariance in the context of assessing the effectiveness of interventions designed to prevent men’s perpetration of IPV.

The present study has two primary objectives to address this knowledge gap. First, employing data from three recent IPV prevention perpetration trials in low- and middle-income countries (LMICs), we aim to explore the factor structure of a scale derived from the Conflict Tactics Scale-Revised (CTS2), the most used measure of men’s perpetration of IPV (Costa and Barros 2016). Second, we rigorously test whether this scale demonstrates measurement invariance across arm, across time, and across studies. Our findings potentially impact the way we interpret and apply results from IPV prevention trials and contribute to the ongoing discourse on the quality and validity of IPV measurement.

## BACKGROUND

The Conflict Tactics Scale (CTS) and its revised, modified, and adapted forms are among the most used scales to assess IPV in community samples (Costa and Barros 2016). The CTS has been used in its original and adapted form in a host of multi-country cross-sectional surveys, such the International Men and Gender Equality Surveys (IMAGES), which has been administered in 32 countries (Equimundo 2022), as well as the United Nations Multi-Country Study on Men and Violence in Asia and the Pacific, which assessed perpetration of IPV and other forms of violence in six countries (Fulu, Jewkes et al. 2013). In its original or adapted forms, the CTS has also been employed as an outcome in evaluations of prevention interventions in settings worldwide (Graham, Embry et al. 2021, DeHond, Brady et al. 2023). The CTS is theoretically grounded in conflict theory, which conceptualizes violence as “an inevitable part of all human association (p.75)” (Straus 1979). The scale measures tactics (18 acts) used by and experienced by each partner in a relationship framed as occurring during a conflict or disagreement. Tactics measured in the CTS include reasoning (later termed negotiation), verbal aggression (later termed psychological aggression), and violence (later termed physical assault), which when tested with principal factors analysis formed three factors (Straus 1979). When two items assessing threats and use of a weapon (knife or gun) were added to the physical assault subscale, Straus reported a fourth factor denoting lethal or “serious” physical assault (1979). However, this factor was comprised of only two items, which is below the minimum recommended threshold of three items to identify a factor (Kenny, Kashy et al. 1998). A review of studies using the CTS and including additional empirical findings (Ballinger III 2000) confirmed the presence in many, but not all studies, of a four-factor solution (reasoning, psychological aggression, physical assault, severe physical assault) and broad similarities in the number of factors for men’s and women’s reports of perpetration. This review also revealed frequent instances of differences in item functioning by gender and a case where the items assessing mild physical assault loaded with psychological aggression for men whereas for women, items measuring psychological aggression and physical assault loaded on separate factors. Also notable were findings of a unidimensional model as the best fit in one study for women given the high correlation among the factors, but for men, both one- and two-factor models had poor fit when only psychological aggression and physical assault were modeled.

In a revised version of the CTS (CTS2), the addition of two subscales capturing sexual coercion and injury from physical assault resulted in five sub-scales (Straus, Hamby et al. 1996). These five factors and to some extent, additional factors denoting levels of severity, have been replicated in some, but not all studies, although most factor analyses have been conducted on samples of women (Jones, Browne et al. 2017). Existing measurement invariance literature involving males has primarily included adolescent and young adult samples. In a study of US college students, a sub-set of psychological and physical IPV perpetration items from the CTS2 demonstrated configural and metric invariance and partial scalar invariance in a two-factor model (Wareham, Wagers et al. 2022). In a study of adolescents in Canada and Italy using a modified version of the CTS2 physical assault sub-scale, invariance of a unidimensional model was found between boys in Canada and Italy and between girls in Canada and Italy, but not between genders within the same country (Nocentini, Menesini et al. 2011). Another study in Chile tested a correlated two-factor model of physical assault (moderate and severe) among college students, which was found to be invariant between men and women (Viejo, Rincón et al. 2018). Finally, examination of a four-factor model (moderate psychological, severe psychological, moderate physical, severe physical) of the Modified-CTS Spanish language version demonstrated gender and age invariance among adolescents and young adults in Northern Spain (Ortuño-Sierra, Marugán Garrido et al. 2023). Collectively, study findings suggest that that a correlated factor structure is the mostly commonly identified model, like that originally proposed by the authors of the CTS, with some possibility of moderate and severe forms of violence forming separate, but correlated constructs. All the studies involving men or adolescent boys were cross-sectional and none included an assessment of sexual violence.

The only measurement invariance study of a CTS-derived scale used in the context of an intervention tested cross-study arm and cross-time invariance using data from four studies of women’s victimization (Clark, Bergenfeld et al. 2023). When assessing the factor structure, three sexual violence items were dropped, two in one study, and one in another due to poor model fit. Confirmatory factor analyses of the remaining items suggested a correlated two-factor model (physical and sexual IPV) was the best fitting model among the three studies that included both physical and sexual IPV. All studies demonstrated strong fit and full invariance by study arm. In invariance analyses over time, all models demonstrated strong fit but only one study demonstrated full invariance. In the remaining three studies, a parameter had to be freed to establish partial invariance including the loading for ‘hit’ in one study, the threshold for ‘hit’ in another study, and the threshold for ‘slap’ in another study; however, these modifications did not meaningfully affect latent means or effect-size estimates.

To date, comparable studies that assess the measurement invariance of measures for men’s IPV perpetration are lacking. We begin to fill this gap by conducting measurement invariance testing using data from three randomized trials of interventions designed to prevent men’s perpetration of IPV. Specifically, we 1) examine the factor structure of the items used to assess men’s perpetration of physical and sexual IPV, 2) conduct tests of measurement invariance across study arms and over time, and 3) assess whether any identified non invariance biases study inferences.

## 2.1 METHODS

### Study Sample

Intervention studies measuring IPV perpetration by men as a primary outcome were identified through the *What Works to Prevent Violence Against Women and Girls Programme* (Crawford, Lloyd-Laney et al. 2020). Investigators participating in *What Works* were encouraged to use standard outcome measures for IPV, but a diversity of study designs and programming types were represented. Three publicly available *What Works* datasets met the present study’s inclusion criteria: an experimental or quasiexperimental panel design with repeated measurement of IPV perpetration at the individual level. All studies were cluster randomized trials that were administered between 2015 and 2019.

Indashyikirwa (IND) was undertaken in rural locations in seven districts in Rwanda among married or cohabitating couples participating in village savings and loans associations. Intervention participants received 21 sessions of group-based training focused on gender, violence, conflict resolution, power dynamics, social norms, economic empowerment, and community change (Stern, Heise et al. 2018). Stepping Stones and Creating Futures (SSCF) was undertaken in urban informal settlements in eThekwini Municipality, KwaZulu-Natal Province, South Africa among men and women 18-30 years old, resident in the community, and not working or in education. Intervention participants received 21 sessions or approximately 63 hours of group education for men and women on gender norms, violence, sexual health, communication and conflict resolution, and livelihood development (Gibbs, Washington et al. 2017). One Man Can (OMC) was deployed in neighborhoods in a semi-formal settlement near Johannesburg, South Africa. The intervention used community mobilization, peer education, and advocacy of varied frequencies to reduce IPV perpetration among community-dwelling men aged 18 to 40 years of age (Christofides, Hatcher et al. 2018).

All studies invited participants to volunteer, and therefore, did not report baseline participation rates. The three datasets originally included 4597 individuals (677 to 1659 across studies) enrolled at baseline. Retention rates ranged from 76% to 97% across studies. Data for the present analysis were subset to include the 3502 men who completed endline data collection (505 to 1537 across studies).

### Data

The *What Works* items measuring physical and sexual IPV perpetration were adapted from the World Health Organization’s (WHO) Multi-country Study on Women’s Health and Domestic Violence Against Women (Garcia-Moreno, Jansen et al. 2005). Adaptations included the combination of two of the severe WHO physical IPV items into one and some additional minor wording adjustments. The WHO scale itself was adapted from items in the Conflicts Tactics Scale (CTS) Revised version (Straus, Hamby et al. 1996, Garcia-Moreno, Jansen et al. 2005).

All studies measured physical IPV in the past 12 months using five items that assessed the frequency with which the respondent did the following to their partners: (1) slapped or threw an object at them; (2) pushed or shoved them; (3) hit them with a fist or object; (4) kicked, dragged, beat, choked, or burnt them; and (5) threatened with or actually attacked them with a weapon (**Table 1**). All studies included at least three items measuring sexually violent acts perpetrated against the respondent’s partner in the past 12 months from among the following: (1) forced to have sex, (2) threatened or intimidated into having sex, (3) forced to do other sexual acts, and (4) forced to view pornography. The IND and OMC studies modified one sexual IPV item each and IND did not ask the question about pornography (**Table 1**). Given the scarcity of data in the higher frequency categories, response options were collapsed to be dichotomous (ever vs never in past 12 months). Items were administered in Kinyarwanda in the IND trial, in English, isi-Zulu, or isiXhosa for the SSCF trial, and in English, isiZulu, Xitsonga or Sepedi in the OMC trial.

**Table 1.** English Language Wording for Items Designed to Measure Physical and Sexual IPV Perpetration, by Study.

| In the past 12 months, how many times have you ... |  | IND | SSCF | OMC |
| --- | --- | --- | --- | --- |
| **SLAP** | slapped a current or previous girlfriend or wife or thrown something at her which could hurt her? | Yes | Yes | Yes |
| **PUSH** | pushed or shoved a current or previous girlfriend or wife? | Yes | Yes | Yes |
| **HIT** | hit a current or previous girlfriend or wife with a fist or with something else which could hurt her? | Yes | Yes | Yes |
| **KICK** | kick, drag, beat, choke, or burn a previous or current girlfriend, partner, or wife? | Yes | Yes | Yes |
| **WEAPON** | threaten to use or actually use a gun, knife, or other weapon against a previous or current girlfriend, partner, or wife? | Yes | Yes | Yes |
| **FORCE SEX** | physically forced your current or previous girlfriend or wife to have sex with you when she did not want to? | Yes | Yes | Yes |
| **COERCED SEX** | used threats or intimidation to get your current or previous partner, girlfriend or wife to have sex when she did not want to? | Yes | Yes | added "but not physical force" |
| **OTHER SEX** | forced your current or previous girlfriend or wife to do something sexual that she did not want to do? | defined forced as "physical force or threats" | Yes | Yes |
| **PORN** | forced your current or previous girlfriend or wife to watch pornography when she didn't want to? | NA | Yes | Yes |
| Notes: Answer options for all three studies included "Never" (0), "One time" (1), "A few times" (2), "Many times" (3). NA = not available. |  |  |  |  |

### Analysis

Univariate analyses of all items were conducted by study arm and time (see **Table 2** for item prevalences). Tetrachoric correlations were estimated to assess the associations among the items at each time point for each study. Average correlations among items also were computed for each study by type of IPV, study arm, and time.

**Table 2.** Percentage Prevalence of Prior-Year Physical and Sexual IPV Items by Study, Study Arm, and Time.

|  | IND (N=1537) |  |  |  | SSCF (N=505) |  |  |  | OMC (N=1460) |  |  |  |
| --- | --- | --- | --- | --- | --- | --- | --- | --- | --- | --- | --- | --- |
|  | Baseline |  | Endline |  | Baseline |  | Endline |  | Baseline |  | Endline |  |
|  | N=763 | N=774 | N=763 | N=774 | N=237 | N=268 | N=237 | N=268 | N=746 | N=714 | N=746 | N=714 |
| ITEM | Trt % | Ctl % | Trt % | Ctl % | Trt % | Ctl % | Trt % | Ctl % | Trt % | Ctl % | Trt % | Ctl % |
| SLAP | 15.0 | 11.5 | 7.0 | 6.8 | 33.3 | 30.7 | 25.3 | 28.4 | 24.0 | 21.8 | 15.1 | 15.6 |
| PUSH | 19.9 | 15.7 | 11.0 | 12.6 | 31.6 | 33.0 | 25.7 | 27.6 | 24.7 | 24.1 | 16.9 | 16.0 |
| HIT | 9.1 | 7.6 | 5.1 | 4.0 | 25.3 | 22.5 | 21.1 | 22.0 | 19.6 | 17.5 | 12.1 | 12.5 |
| KICK | 2.2 | 3.2 | 1.6 | 1.2 | 18.6 | 17.6 | 15.6 | 19.4 | 16.2 | 13.5 | 10.7 | 11.4 |
| WEAPON | 0.9 | 1.8 | 1.3 | 0.9 | 13.5 | 11.2 | 13.9 | 13.8 | 11.9 | 11.3 | 6.7 | 9.7 |
| FORCED SEX | 13.1 | 11.1 | 5.8 | 7.4 | 16.9 | 18.7 | 15.2 | 17.2 | 17.0 | 15.2 | 10.6 | 11.0 |
| COERCED SEX | 17.2 | 15.5 | 8.5 | 12.8 | 19.0 | 17.6 | 12.7 | 16.0 | 15.8 | 15.4 | 10.1 | 9.4 |
| OTHER SEX | 2.5 | 3.1 | 2.6 | 2.5 | 16.9 | 20.6 | 15.2 | 19.8 | 18.0 | 16.1 | 9.8 | 10.5 |
| PORN | na | na | na | na | 14.8 | 15.5 | 14.8 | 17.2 | 15.7 | 14.0 | 9.8 | 11.0 |
| **Notes:** IND: Indashyikirwa; SSCF: Stepping Stones and Creating Futures; OMC: One Man Can. |  |  |  |  |  |  |  |  |  |  |  |  |

To assess the factor structure of the physical and sexual IPV items, we implemented incremental exploratory factor analyses (EFA) of one to two factors, which is the largest number of theoretically relevant factors across studies that could be fit, given the small number of items available for analysis. These models utilized a random split-half sample and accounted for the complex sampling design of each study. Model fit was assessed using well-established metrics and cut points of good fit, including the Root Mean Square Error of Approximation (RMSEA <0.06), Comparative Fit Index, and Tucker-Lewis Index (CFI, TLI >0.95) (Hu and Bentler 1995, Hu and Bentler 1999, Vandenberg and Lance 2000, Cheung and Rensvold 2002, Putnick and Bornstein 2016). Using the best fitting model for each study, we fit models using confirmatory factor analysis (CFA) in the second random split half of the samples. Given prior research demonstrating the likelihood that a bifactor model may be the best fitting model (Ballinger III 2000), we also implemented a bifactor analysis of the study with the largest sample size.

We performed measurement invariance testing using multiple group confirmatory factor analysis (MGCFA) for dichotomous indicators (Millsap 2012). We compared the fit of models with and without factor loadings and thresholds constrained (Davidov, Datler et al. 2012). Model fit was assessed using the criteria described above with non-invariance being defined by a worsening in RMSEA and CFI of more than 0.01 (Liu, Millsap et al. 2017).

For items showing any non-invariance, we used maximum likelihood estimation to determine whether non-invariance arose primarily from loadings or thresholds. Where there was a lack of support for invariance through comparison of configural versus metric models, modification indices from weighted least square mean and variance adjusted (WLSMV) estimation were used to identify potential constraints to relax. We had planned to estimate the severity of the impact of non-invariance on cross-arm comparisons by estimating the standardized differences of latent IPV means between models with and without equality restrictions and calculate the change in the difference-in-difference estimates as a proportion of the standard deviation. However, as will be seen below, one of the studies demonstrated scalar invariance, not requiring further testing, one achieved only configural invariance, precluding further analysis, and one achieved scalar invariance in post hoc analyses. For this study, the analytic approach described in its impact paper was replicated using the invariant form of the IPV perpetration variable. The reported impact analysis was a cluster-level comparison of endline perpetration scores, adjusting for baseline perpetration and unbalanced baseline covariates and weighting with inverse probability to treatment weights to account for loss-to-follow up (Christofides, Hatcher et al. 2020).

## RESULTS

Prevalence of physical IPV in the prior 12-months ranged from 24.3% (IND) to 48.0% (SSCF) at baseline and 15.6% (IND) to 40.4% (SSCF) at endline (Table 2). Slap and push were the most frequently reported acts of physical IPV, and weapon was the least frequently reported act across studies and time. The prevalence of sexual IPV in the prior 12-months ranged from 21.0% (IND) to 32.3% (SSCF) at baseline and 13.7% (IND) to 27.7% (SSCF) at endline, with greater variability in the most and least frequently reported items across studies and time.

**Table 3** displays the tetrachoric correlations among the items by IPV type, study arm, and assessment wave for each study. The average correlation among items measuring physical IPV ranged from 0.68 to 0.73 across the studies at baseline and 0.72 to 0.88 at endline. The average correlation among items measuring sexual IPV ranged between 0.54 to 0.75 across the studies at baseline and rose to a range of 0.58 to 0.93 at endline. The average correlation among all items followed a similar increasing trend over time. The average increase in correlation among physical IPV items ranged from 0.03 (SSCF) to 0.15 (OMC). These increases were in the treatment arm in all studies, although the OMC trial also saw equally large increases in the correlation among these items in the control arm. The average correlation among sexual IPV items ranged from 0.07 (IND) to 0.17 (OMC). These increases were influenced by the treatment arm in the IND trial, the control arm in the SSCF trial, and both arms in the OMC trial. Across studies, the correlation of items within IPV types was generally higher than across types. In the SSCF and OMC trials, several items correlated with other items ≥ 0.85 within the same type for physical and sexual IPV at endline (SSCF: ‘kick,’ ‘coerced sex’, ‘other sex’), with all items showing this trend in the OMC trial including most sexual abuse items, which correlated over 0.90. Item-level correlations for each study are available in the online supplement.

**Table 3.** Correlations Among Items by Study, IPV Type, Study Arm, and Time.

|  | Baseline |  |  | Endline |  |  | Difference<br>(Endline – Baseline) |  |  |
| --- | --- | --- | --- | --- | --- | --- | --- | --- | --- |
|  | Mean | Tx | Ctl | Mean | Tx | Ctl | Mean | Tx | Ctl |
| IND: Physical | 0.68 | 0.53 | 0.76 | 0.72 | 0.74 | 0.69 | 0.04 | 0.21 | -0.07 |
| IND: Sexual | 0.54 | 0.43 | 0.63 | 0.61 | 0.58 | 0.63 | 0.07 | 0.16 | 0.00 |
| IND: Physical and Sexual | 0.55 | 0.40 | 0.63 | 0.61 | 0.62 | 0.60 | 0.07 | 0.22 | -0.03 |
| SSCF: Physical | 0.73 | 0.72 | 0.74 | 0.76 | 0.81 | 0.71 | 0.03 | 0.09 | -0.03 |
| SSCF: Sexual | 0.73 | 0.83 | 0.63 | 0.82 | 0.84 | 0.80 | 0.09 | 0.01 | 0.17 |
| SSCF: Physical and Sexual | 0.65 | 0.69 | 0.61 | 0.70 | 0.75 | 0.66 | 0.05 | 0.05 | 0.05 |
| OMC: Physical | 0.73 | 0.74 | 0.73 | 0.88 | 0.88 | 0.89 | 0.15 | 0.14 | 0.16 |
| OMC: Sexual | 0.75 | 0.79 | 0.70 | 0.93 | 0.93 | 0.92 | 0.17 | 0.14 | 0.22 |
| OMC: Physical and Sexual | 0.66 | 0.67 | 0.65 | 0.81 | 0.82 | 0.80 | 0.15 | 0.15 | 0.16 |
| **Notes:** IND: Indashyikirwa; SSCF: Stepping Stones and Creating Futures; OMC: One Man Can. Tx: Treatment; Ctl: Control. |  |  |  |  |  |  |  |  |  |

In the EFA, the best fitting model was a two-factor solution for all studies (**Table 4**). Of the two-factor models, the OMC study was the ‘cleanest’ with no cross-loadings and all physical IPV items loaded on a factor together while all sexual IPV items loaded together on the other factor. The SSCF model had two cross loadings (‘weapon’ and ‘porn’) while the IND study had one cross loading (‘weapon’). The EFAs with item-level detail are available in the online supplement.

**Table 4.**
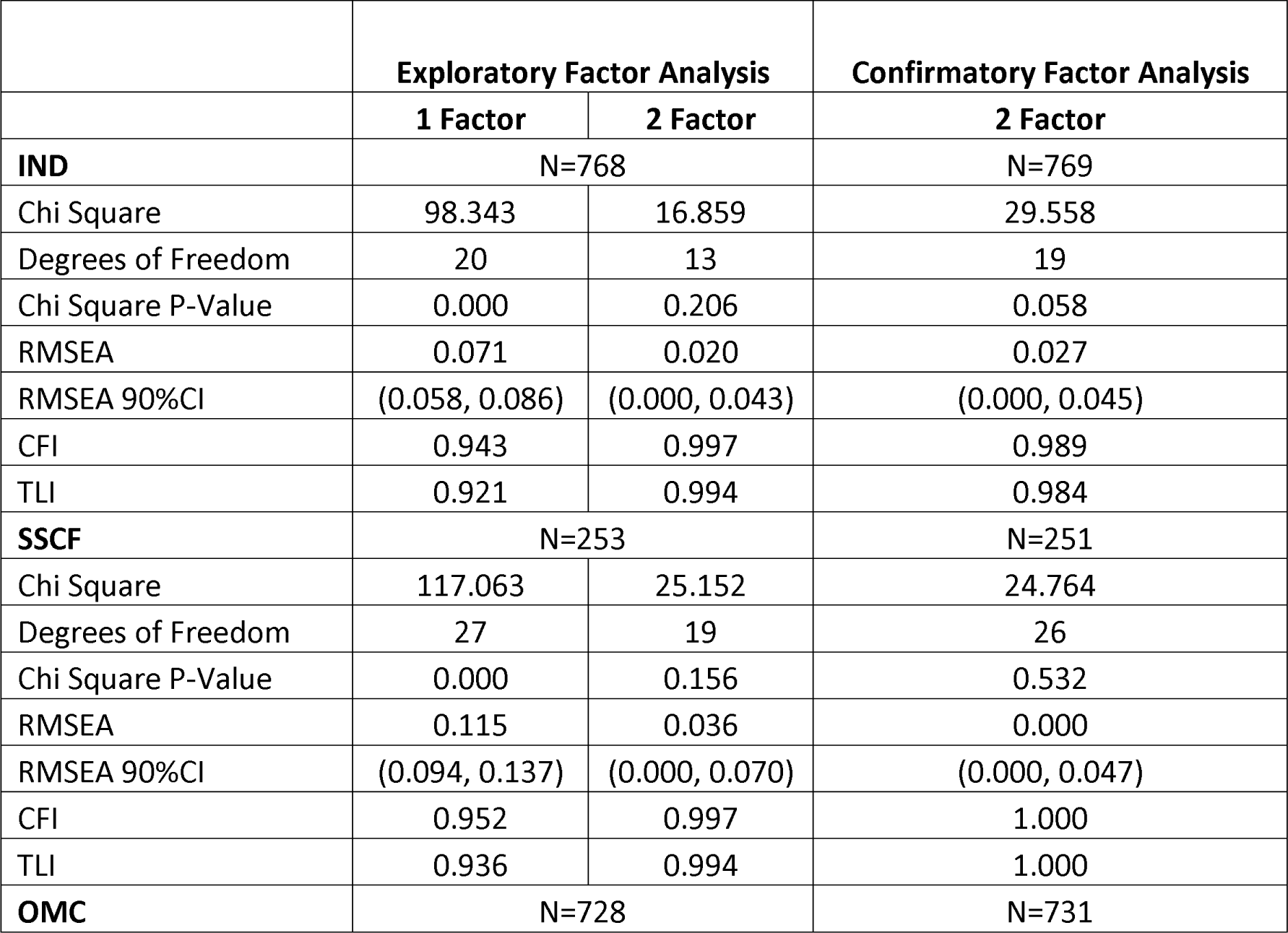
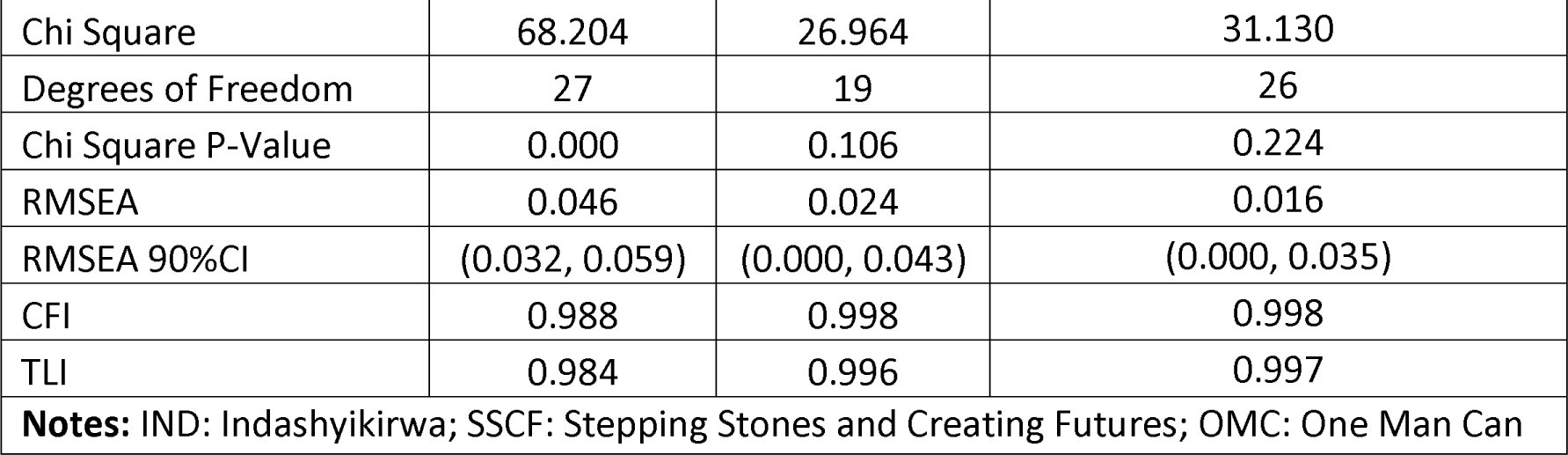
Exploratory and Confirmatory Factor Analysis by Study, Baseline.

With such a limited item set and on theoretical grounds we opted not to drop cross-loading items from their respective studies but to proceed with the theoretically aligned two-factor CFAs. Across all studies, the two-factor CFAs had very strong fit (Table 4). Chi-square difference testing between a one- and a two-factor solution confirmed this finding. The CFAs with item-level detail are available in the online supplement. We also tested whether a bifactor model would be a stronger fit than a two-factor model in the largest study (IND) given prior CTS findings to this effect. We fit an exploratory bifactor model on a random split half of the sample and a confirmatory bifactor model on the remaining split half. While the confirmatory bifactor model fit the data well (RMSEA: 0.04; CFI: 0.98; TLI: 0.96) the average parameter bias was 0.22, suggesting that the bifactor results were not trustworthy.

The two-factor model for each study was subjected to invariance testing by study arm and time (see **Table 5** for summary, see **Table 6** for detailed results).

**Table 5.**
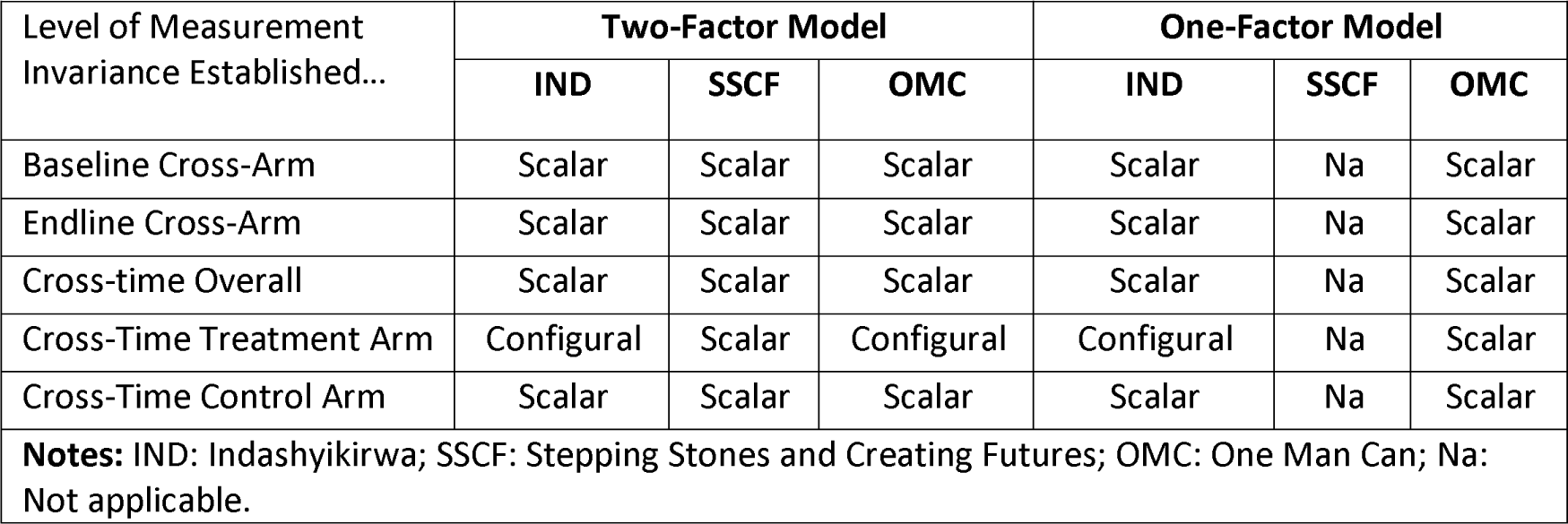
Measurement Invariance Testing Summary by Study.

**Table 6.**
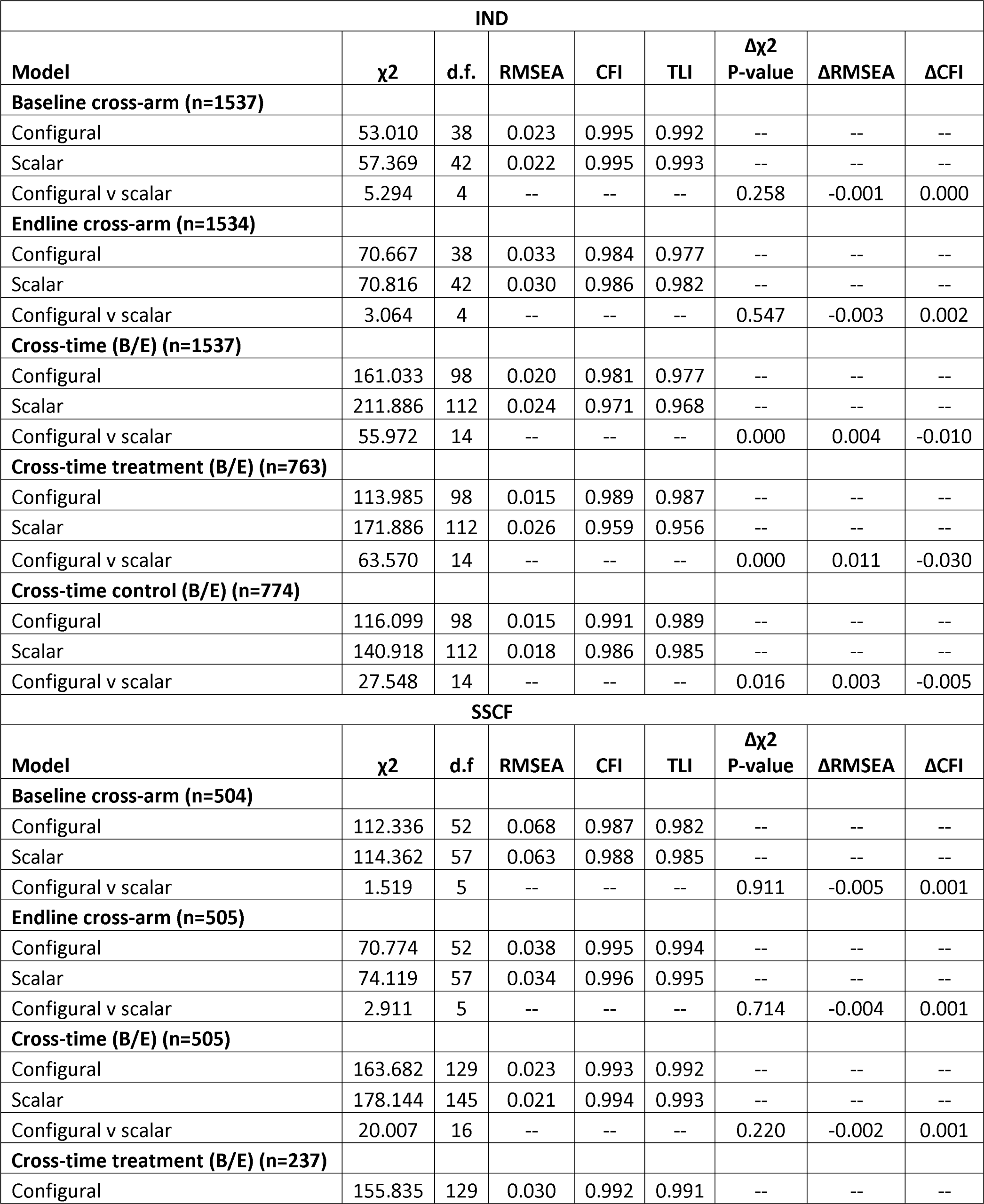
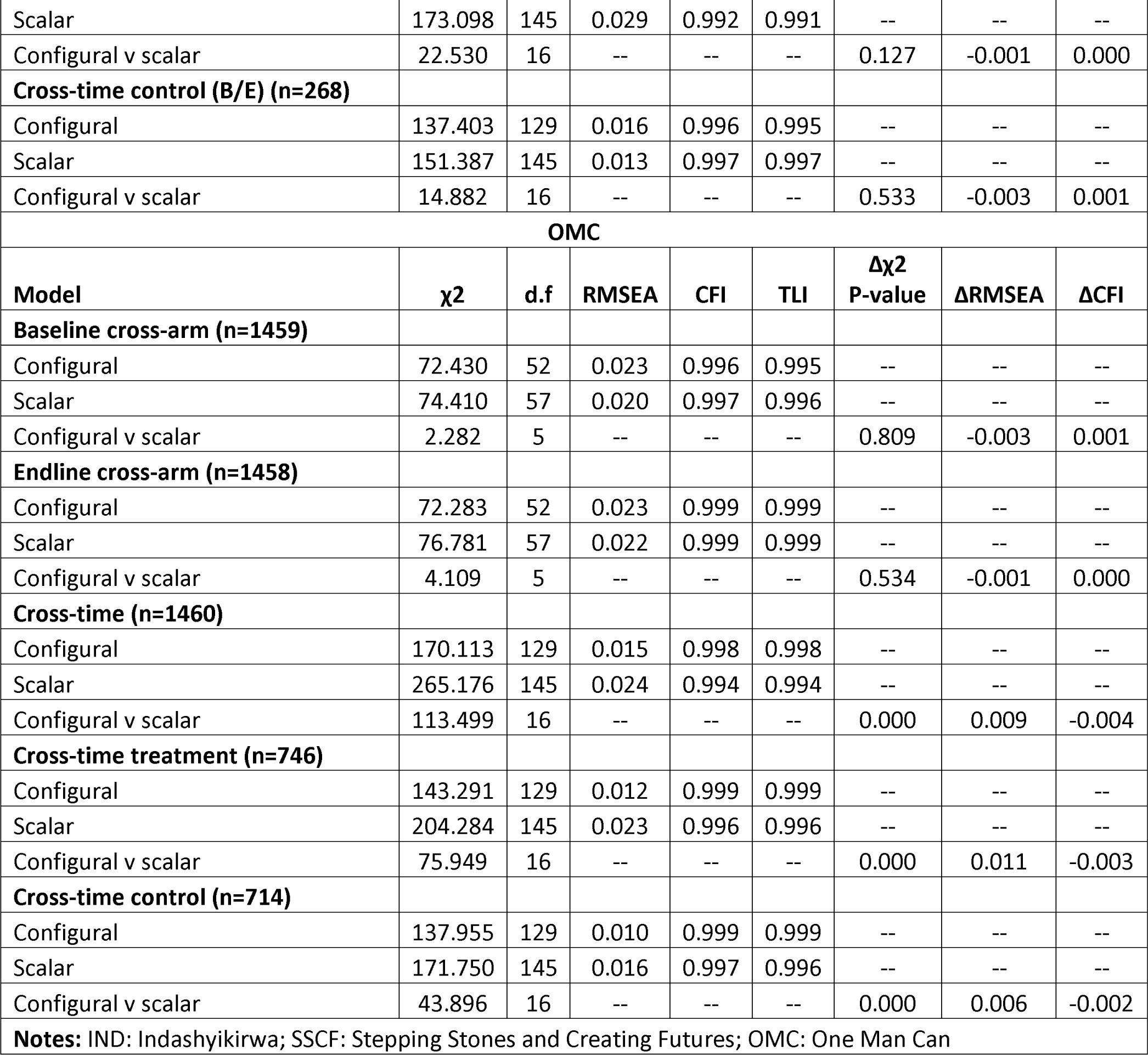
Measurement Invariance Testing by Study.

| IND |  |  |  |  |  |  |  |  |
| --- | --- | --- | --- | --- | --- | --- | --- | --- |
| Model | $\chi^2$ | d.f. | RMSEA | CFI | TLI | $\Delta\chi^2$<br>P-value | ΔRMSEA | ΔCFI |
| **Baseline cross-arm (n=1537)** |  |  |  |  |  |  |  |  |
| Configural | 53.010 | 38 | 0.023 | 0.995 | 0.992 | -- | -- | -- |
| Scalar | 57.369 | 42 | 0.022 | 0.995 | 0.993 | -- | -- | -- |
| Configural v scalar | 5.294 | 4 | -- | -- | -- | 0.258 | -0.001 | 0.000 |
| **Endline cross-arm (n=1534)** |  |  |  |  |  |  |  |  |
| Configural | 70.667 | 38 | 0.033 | 0.984 | 0.977 | -- | -- | -- |
| Scalar | 70.816 | 42 | 0.030 | 0.986 | 0.982 | -- | -- | -- |
| Configural v scalar | 3.064 | 4 | -- | -- | -- | 0.547 | -0.003 | 0.002 |
| **Cross-time (B/E) (n=1537)** |  |  |  |  |  |  |  |  |
| Configural | 161.033 | 98 | 0.020 | 0.981 | 0.977 | -- | -- | -- |
| Scalar | 211.886 | 112 | 0.024 | 0.971 | 0.968 | -- | -- | -- |
| Configural v scalar | 55.972 | 14 | -- | -- | -- | 0.000 | 0.004 | -0.010 |
| **Cross-time treatment (B/E) (n=763)** |  |  |  |  |  |  |  |  |
| Configural | 113.985 | 98 | 0.015 | 0.989 | 0.987 | -- | -- | -- |
| Scalar | 171.886 | 112 | 0.026 | 0.959 | 0.956 | -- | -- | -- |
| Configural v scalar | 63.570 | 14 | -- | -- | -- | 0.000 | 0.011 | -0.030 |
| **Cross-time control (B/E) (n=774)** |  |  |  |  |  |  |  |  |
| Configural | 116.099 | 98 | 0.015 | 0.991 | 0.989 | -- | -- | -- |
| Scalar | 140.918 | 112 | 0.018 | 0.986 | 0.985 | -- | -- | -- |
| Configural v scalar | 27.548 | 14 | -- | -- | -- | 0.016 | 0.003 | -0.005 |
| SSCF |  |  |  |  |  |  |  |  |
| Model | $\chi^2$ | d.f. | RMSEA | CFI | TLI | $\Delta\chi^2$<br>P-value | ΔRMSEA | ΔCFI |
| **Baseline cross-arm (n=504)** |  |  |  |  |  |  |  |  |
| Configural | 112.336 | 52 | 0.068 | 0.987 | 0.982 | -- | -- | -- |
| Scalar | 114.362 | 57 | 0.063 | 0.988 | 0.985 | -- | -- | -- |
| Configural v scalar | 1.519 | 5 | -- | -- | -- | 0.911 | -0.005 | 0.001 |
| **Endline cross-arm (n=505)** |  |  |  |  |  |  |  |  |
| Configural | 70.774 | 52 | 0.038 | 0.995 | 0.994 | -- | -- | -- |
| Scalar | 74.119 | 57 | 0.034 | 0.996 | 0.995 | -- | -- | -- |
| Configural v scalar | 2.911 | 5 | -- | -- | -- | 0.714 | -0.004 | 0.001 |
| **Cross-time (B/E) (n=505)** |  |  |  |  |  |  |  |  |
| Configural | 163.682 | 129 | 0.023 | 0.993 | 0.992 | -- | -- | -- |
| Scalar | 178.144 | 145 | 0.021 | 0.994 | 0.993 | -- | -- | -- |
| Configural v scalar | 20.007 | 16 | -- | -- | -- | 0.220 | -0.002 | 0.001 |
| **Cross-time treatment (B/E) (n=237)** |  |  |  |  |  |  |  |  |
| Configural | 155.835 | 129 | 0.030 | 0.992 | 0.991 | -- | -- | -- |
| Scalar | 173.098 | 145 | 0.029 | 0.992 | 0.991 | -- | -- | -- |
| Configural v scalar | 22.530 | 16 | -- | -- | -- | 0.127 | -0.001 | 0.000 |
| **Cross-time control (B/E) (n=268)** |  |  |  |  |  |  |  |  |
| Configural | 137.403 | 129 | 0.016 | 0.996 | 0.995 | -- | -- | -- |
| Scalar | 151.387 | 145 | 0.013 | 0.997 | 0.997 | -- | -- | -- |
| Configural v scalar | 14.882 | 16 | -- | -- | -- | 0.533 | -0.003 | 0.001 |
| **OMC** |  |  |  |  |  |  |  |  |
| **Model** | **$\chi^2$** | **d.f** | **RMSEA** | **CFI** | **TLI** | **$\Delta\chi^2$**<br>**P-value** | **ΔRMSEA** | **ΔCFI** |
| **Baseline cross-arm (n=1459)** |  |  |  |  |  |  |  |  |
| Configural | 72.430 | 52 | 0.023 | 0.996 | 0.995 | -- | -- | -- |
| Scalar | 74.410 | 57 | 0.020 | 0.997 | 0.996 | -- | -- | -- |
| Configural v scalar | 2.282 | 5 | -- | -- | -- | 0.809 | -0.003 | 0.001 |
| **Endline cross-arm (n=1458)** |  |  |  |  |  |  |  |  |
| Configural | 72.283 | 52 | 0.023 | 0.999 | 0.999 | -- | -- | -- |
| Scalar | 76.781 | 57 | 0.022 | 0.999 | 0.999 | -- | -- | -- |
| Configural v scalar | 4.109 | 5 | -- | -- | -- | 0.534 | -0.001 | 0.000 |
| **Cross-time (n=1460)** |  |  |  |  |  |  |  |  |
| Configural | 170.113 | 129 | 0.015 | 0.998 | 0.998 | -- | -- | -- |
| Scalar | 265.176 | 145 | 0.024 | 0.994 | 0.994 | -- | -- | -- |
| Configural v scalar | 113.499 | 16 | -- | -- | -- | 0.000 | 0.009 | -0.004 |
| **Cross-time treatment (n=746)** |  |  |  |  |  |  |  |  |
| Configural | 143.291 | 129 | 0.012 | 0.999 | 0.999 | -- | -- | -- |
| Scalar | 204.284 | 145 | 0.023 | 0.996 | 0.996 | -- | -- | -- |
| Configural v scalar | 75.949 | 16 | -- | -- | -- | 0.000 | 0.011 | -0.003 |
| **Cross-time control (n=714)** |  |  |  |  |  |  |  |  |
| Configural | 137.955 | 129 | 0.010 | 0.999 | 0.999 | -- | -- | -- |
| Scalar | 171.750 | 145 | 0.016 | 0.997 | 0.996 | -- | -- | -- |
| Configural v scalar | 43.896 | 16 | -- | -- | -- | 0.000 | 0.006 | -0.002 |
| **Notes:** IND: Indashyikirwa; SSCF: Stepping Stones and Creating Futures; OMC: One Man Can |  |  |  |  |  |  |  |  |

The baseline and endline cross-study arm tests all demonstrated scalar invariance, suggesting that the scale had similar measurement properties across arms at these two time points. The SSCF trial demonstrated invariance across time overall and within each study arm. Both the IND and OMC trials demonstrated scalar non-invariance in the intervention arms over time, suggesting that the measurement properties of the perpetration scale differed over time in the intervention arms. In the maximum likelihood models for each study, the invariance was concentrated in the thresholds denoted by the likelihood ratio tests comparing 1) the configural to the metric model and 2) the metric to the scalar models. In the OMC trial, there were no suggested threshold or loading modifications that would improve the model fit. In the IND trial, the ‘slap’ threshold was the only threshold identified along with its loading. Freeing the threshold in subsequent models in an attempt to establish partial scalar invariance did not result in adequate model fit, nor did freeing both its threshold and factor loading and no other loadings or thresholds were identified as modifications that would improve model fit. In post hoc testing of a more parsimonious one-factor model, the OMC study achieved measurement invariance across study arms and time (ΔRMSEA and ΔCFI <0.01 for all models); the IND trial did not. The measurement of IPV was noninvariant over time in the intervention arm (ΔRMSEA: 0.00 and ΔCFI: −0.026). In further post hoc testing on the IND trial data to attempt to reach partial scalar invariance, we made several other modifications including dropping the ‘coerced sex’ item in a one-factor model due to its very high correlation with ‘forced sex’, dropping the ‘other sex’ item due it having the lowest loading in the EFA model, and adding a correlation among the ‘forced sex’ and ‘coerced sex’ items at endline and at both baseline and endline in one and two factor models. The results were the same across all attempts, namely evidence of scalar non-invariance in the intervention arm over time (see the online supplement for results).

### Concordance with Published Study Results

Given the invariance in the SSCF trial no re-examination of trial results was needed. The inability to identify a measurement model in the IND trial that was invariant precluded further analysis, as the scale is not measuring the same construct over time in the treatment and control arms. For the OMC trial, we replicated the trial’s endline analysis using the one-factor invariant model finding similar null results (estimate 0.02, p-value 0.42).

## DISCUSSION

The CTS2, and scales derived from it, are the most widely used measures of adult men’s IPV perpetration globally for surveillance and impact assessments but remain untested for their valid use to determine the effectiveness of interventions. This novel study begins to address this gap, finding that the scale cannot be validly used to assess intervention impact without measurement invariance testing demonstrating at least partial scalar invariance across arms and study waves in the trial. Unlike results of a similar measurement invariance study of women IPV’s victimization (Clark, Bergenfeld et al. 2023), where at least partial scalar invariance was found within and across trials and the identified non-invariance had negligible impact on inferences regarding intervention effects, the findings among men suggest considerable caution is needed. Only one of the studies demonstrated measurement invariance across study arms and time to enable valid inference. One study only demonstrated measurement invariance after considerable post hoc testing and when modeled differently than the trial findings, and one never attained measurement invariance in the intervention arm over time, precluding comparison of change in IPV perpetration between the intervention and control groups, which is foundational to assessing the impact of a prevention intervention.

Across trials, the best fitting exploratory and confirmatory model, a correlated two-factor solution, is aligned with prior research on the CTS (Straus 1979, Straus, Hamby et al. 1996, Ballinger III 2000, Jones, Browne et al. 2017). However, this model was invariant in only one trial (SSCF), with a more parsimonious one-factor solution needed for invariance in another trial (OMC). In this trial (OMC), the high correlations within physical and sexual IPV might have driven the performance of the one-factor model. However, the very high correlation among items across studies, especially for sexual IPV suggest that respondents may not differentiating among the items, particularly as the items are similarly worded. The likely overlap in the domain being measured suggests inefficient measurement and a potential lack of content validity.

The increasing correlation among the items over time, especially in the IND trial and the concentration of measurement invariance in the treatment arms in the IND and OMC trials suggests that treatment may have an impact on the nature of the construct over time. Changes in the correlations might reflect the impact of the intervention such that individuals exposed to the intervention may have changed the way they interpreted the questions, or they can reflect other methodological issues such as homogenization of responses, common experience effect, or social desirability bias. However, the OMC trial, the only trial to attempt exposure among community members instead of focused intervention on individual men, reported limited intervention exposure, suggesting that factors other than the treatment are likely affecting measurement over time. For example, the participants may have experienced other unanticipated exposures in the treatment and control arms, given the change in correlation among both arms.

The implication of this study’s findings on published reports of trial outcomes suggests that considerable caution is needed when using this scale to measure intervention impact since it was a valid measure of impact in only one trial. For the trial in which the correlated two-factor solution was invariant (SSCF), study findings support the validity of the a reduction in men’s reports of perpetration for those in the intervention compared to the control arm (Gibbs, Washington et al. 2020). For the OMC trial, the use of a single factor form of the outcome variable (the only form that was invariant) did not appreciably alter the study’s null finding (Christofides, Hatcher et al. 2020). The difference between the treatment and control arm was so small that using an invariant form of the outcome variable made no difference. For the IND trial, the reported significant reduction in men’s reports of perpetration compared to the control arm (Dunkle, Stern et al. 2020) does not appear to be valid given the lack of measurement invariance in the treatment arm over time. However, the IND trial also reported a reduction in women’s reports of IPV victimization, and the measure was found to be partially invariant (threshold for slap had to be freed to establish invariance over time) but this minor modification did not affect study inference (Clark, Bergenfeld et al. 2023). If there had been no measurement of women’s victimization, the implications of this study’s findings on trial inference would potentially be severe.

### Limitations and Strengths

The findings must be considered in light of study limitations. We have relied on the English translations of the items used in the studies. The surveys were administrated in up to four different languages and the underlying quality of the translations is unknown. The one study in which invariance was detected also had the smallest sample size and therefore the most limited power. However, the study relied on alternative fit indices (RMSEA, CFI, TLI) for model fit determination, which are less sensitive to sample size (Cheung and Rensvold 2002). The small number of studies and limited geographic scope limits generalizability of our findings beyond these studies. Despite these limitations, the studies were diverse in terms of preventive intervention programming and utilized among the most common items in LMICs for surveillance and evaluations, providing a strong basis to begin to assess the utility of these items for impact assessments.

### Research and Policy Implications

As has been documented among invariance tests of women’s reports of IPV victimization in intervention trials (Clark, Bergenfeld et al. 2023) and national surveys (Yount, Bergenfeld et al. 2022, Yount, Cheong et al. 2022), there are several lessons to be drawn, which may be even more important for measuring men’s IPV perpetration. First, there is a lack of consensus on the definition and scope of the construct of IPV perpetration. The basis of the scales used in this study were developed in the 1970s and revised in the 1990s to measure relationship conflict in high-income settings in the Global North. Despite the CTS being designed to measure men’s and women’s victimization and perpetration, the findings from this study suggest that the scale performs more poorly among men than prior research among women using identical or nearly identical items (Clark, Bergenfeld et al. 2023) warranting caution when using the item set to assess the impact of an intervention among men.

Second, despite an attempt by the *What Works* Consortium to standardize measurement of IPV across studies, in some studies, items were modified. While pooled testing was not warranted due to variation in factor structures across studies, the items were designed to measure the same act. Modifications in the items increase the likelihood that they are interpreted differently across the studies leading to non-invariance. The push to modify the sexual IPV items is likely due, in part, to their lack of specificity, which allows for a very large range of interpretations, the opposite of what a survey item is designed to elicit.

Third, the small number of items limits the investigator’s ability to drop items should they show item-specific non-invariance across arms and time. While very large item sets are impractical, the current very small item set leaves little flexibility for dropping non-invariance items. The small item set also limits an assessment of severity, as prior research on the CTS and CTS2 are suggestive of separate factors by levels of severity, which if assessed would offer additional insights into the impact of interventions across these sub-domains. The limited number of items and very high correlations also suggest inefficient measurement with limited content validity.

Fourth, in the short-term, further measurement-focused research on a larger number of studies across geographic settings and intervention types is needed to ascertain the extent of the problem of non-invariance and whether certain intervention content causes cognitive shifts in interpretation or propensity to report IPV perpetration. Research is also needed that eventually leads to the development of scales that can be validly compared across groups, studies, and time and relied upon without further measurement invariance testing.

Finally, the lack of invariance across the trials has implications for policy makers. Evidence-based decision-making requires confidence in study findings. The lack of consistency in the performance of the perpetration items suggests that caution is needed when identifying effective interventions for replication and scale up and has knock-on effects on the veracity of cost-effectiveness assessments, critical considerations when choosing how to invest scarce resources.

## CONCLUSION

Taken together the findings suggest that the CTS cannot be used to make valid comparisons of intervention effects without further refinement. Until such time, however, measurement invariance testing is needed to ensure valid study inferences are drawn from IPV perpetration trials when the CTS2 item set is the outcome of interest.

## Data Availability

The trial data used in this manuscript are publicly at the MEDAT Data Repository - SAMRC

http://medat.samrc.ac.za/index.php/catalog/WW

## Author Contributions

Cari J. Clark: Conceptualization, Methodology, Formal analysis, Visualization, Resources, Supervision, Project Administration, Writing - original draft, Writing - review and editing, Funding acquisition

Irina Bergenfeld: Data curation, Formal analysis, Visualization, Writing - original draft, Writing - review & editing

Abbie Shervinskie: Data curation, Formal analysis, Visualization, Writing - original draft, Writing - review & editing

Erin R. Johnson: Data curation, Formal analysis, Visualization, Writing - original draft, Writing - review & editing

Yuk Fai Cheong: Conceptualization, Methodology, Resources, Supervision, Writing - review & editing; Funding acquisition

Nadine J. Kaslow: Writing-review & editing

Kathryn M. Yount: Conceptualization, Methodology, Resources, Supervision, Project Administration, Writing - review and editing, Funding acquisition

