## Supplement for "Validity of a Common Measure of Intimate Partner Violence Perpetration: Impact on Study Inference in Trials in Low- and Middle-Income Countries"

**Online Supplement Table 1. Item-Level Correlation by Time and Study.**

| **IND (N=1537)** | **SLAP0** | **PUSH0** | **HIT0** | **KICK0** | **WEAPON0** | **FORCE0** | **COERCE0** | **OTHSEX0** | **PORN0** | **SLAP2** | **PUSH2** | **HIT2** | **KICK2** | **WEAPON2** | **FORCE2** | **COERCE2** | **OTHSEX2** | **PORN2** |
| --- | --- | --- | --- | --- | --- | --- | --- | --- | --- | --- | --- | --- | --- | --- | --- | --- | --- | --- |
| **SLAP0** |  |  |  |  |  |  |  |  |  |  |  |  |  |  |  |  |  |  |
| **PUSH0** | 0.74 |  |  |  |  |  |  |  |  |  |  |  |  |  |  |  |  |  |
| **HIT0** | 0.81 | 0.74 |  |  |  |  |  |  |  |  |  |  |  |  |  |  |  |  |
| **KICK0** | 0.62 | 0.65 | 0.77 |  |  |  |  |  |  |  |  |  |  |  |  |  |  |  |
| **WEAPON0** | 0.54 | 0.55 | 0.67 | 0.77 |  |  |  |  |  |  |  |  |  |  |  |  |  |  |
| **FORCE0** | 0.50 | 0.48 | 0.52 | 0.57 | 0.67 |  |  |  |  |  |  |  |  |  |  |  |  |  |
| **COERCE0** | 0.46 | 0.47 | 0.45 | 0.50 | 0.49 | 0.84 |  |  |  |  |  |  |  |  |  |  |  |  |
| **OTHSEX0** | 0.24 | 0.21 | 0.30 | 0.45 | 0.50 | 0.35 | 0.44 |  |  |  |  |  |  |  |  |  |  |  |
| **PORN0** | NA | NA | NA | NA | NA | NA | NA | NA |  |  |  |  |  |  |  |  |  |  |
| **SLAP2** | 0.48 | 0.38 | 0.39 | 0.45 | 0.37 | 0.37 | 0.26 | 0.23 | NA |  |  |  |  |  |  |  |  |  |
| **PUSH2** | 0.53 | 0.49 | 0.46 | 0.44 | 0.56 | 0.34 | 0.34 | 0.20 | NA | 0.73 |  |  |  |  |  |  |  |  |
| **HIT2** | 0.44 | 0.41 | 0.45 | 0.42 | 0.46 | 0.48 | 0.37 | 0.23 | NA | 0.81 | 0.74 |  |  |  |  |  |  |  |
| **KICK2** | 0.37 | 0.30 | 0.52 | 0.62 | 0.70 | 0.30 | 0.16 | 0.43 | NA | 0.71 | 0.85 | 0.81 |  |  |  |  |  |  |
| **WEAPON2** | 0.29 | 0.08 | 0.08 | 0.30 | 0.26 | 0.09 | -0.21 | 0.01 | NA | 0.59 | 0.57 | 0.62 | 0.83 |  |  |  |  |  |
| **FORCE2** | 0.28 | 0.24 | 0.35 | 0.33 | 0.44 | 0.57 | 0.46 | 0.28 | NA | 0.51 | 0.56 | 0.65 | 0.75 | 0.60 |  |  |  |  |
| **COERCE2** | 0.30 | 0.19 | 0.27 | 0.42 | 0.27 | 0.47 | 0.47 | 0.23 | NA | 0.47 | 0.46 | 0.56 | 0.66 | 0.45 | 0.87 |  |  |  |
| **OTHSEX2** | 0.20 | 0.12 | 0.25 | -0.01 | 0.27 | 0.23 | 0.14 | 0.30 | NA | 0.44 | 0.27 | 0.47 | 0.58 | 0.68 | 0.51 | 0.45 |  |  |
| **PORN2** | NA | NA | NA | NA | NA | NA | NA | NA | NA | NA | NA | NA | NA | NA | NA | NA | NA |  |
| **OMC (N=1460)** | **SLAP0** | **PUSH0** | **HIT0** | **KICK0** | **WEAPON0** | **FORCE0** | **COERCE0** | **OTHSEX0** | **PORN0** | **SLAP2** | **PUSH2** | **HIT2** | **KICK2** | **WEAPON2** | **FORCE2** | **COERCE2** | **OTHSEX2** | **PORN2** |
| **SLAP0** |  |  |  |  |  |  |  |  |  |  |  |  |  |  |  |  |  |  |
| **PUSH0** | 0.74 |  |  |  |  |  |  |  |  |  |  |  |  |  |  |  |  |  |
| **HIT0** | 0.76 | 0.73 |  |  |  |  |  |  |  |  |  |  |  |  |  |  |  |  |
| **KICK0** | 0.71 | 0.74 | 0.79 |  |  |  |  |  |  |  |  |  |  |  |  |  |  |  |
| **WEAPON0** | 0.65 | 0.67 | 0.74 | 0.80 |  |  |  |  |  |  |  |  |  |  |  |  |  |  |
| **FORCE0** | 0.50 | 0.55 | 0.58 | 0.57 | 0.58 |  |  |  |  |  |  |  |  |  |  |  |  |  |
| **COERCE0** | 0.57 | 0.57 | 0.65 | 0.63 | 0.65 | 0.81 |  |  |  |  |  |  |  |  |  |  |  |  |
| **OTHSEX0** | 0.59 | 0.59 | 0.69 | 0.68 | 0.68 | 0.76 | 0.80 |  |  |  |  |  |  |  |  |  |  |  |
| **PORN0** | 0.46 | 0.55 | 0.61 | 0.64 | 0.65 | 0.66 | 0.70 | 0.77 |  |  |  |  |  |  |  |  |  |  |
| **SLAP2** | 0.30 | 0.29 | 0.27 | 0.35 | 0.25 | 0.30 | 0.19 | 0.25 | 0.27 |  |  |  |  |  |  |  |  |  |
| **PUSH2** | 0.30 | 0.34 | 0.33 | 0.32 | 0.27 | 0.28 | 0.24 | 0.29 | 0.32 | 0.89 |  |  |  |  |  |  |  |  |
| **HIT2** | 0.28 | 0.32 | 0.37 | 0.39 | 0.27 | 0.28 | 0.22 | 0.26 | 0.30 | 0.92 | 0.86 |  |  |  |  |  |  |  |
| **KICK2** | 0.20 | 0.27 | 0.30 | 0.39 | 0.26 | 0.25 | 0.16 | 0.24 | 0.31 | 0.89 | 0.84 | 0.92 |  |  |  |  |  |  |
| **WEAPON2** | 0.21 | 0.34 | 0.34 | 0.37 | 0.29 | 0.23 | 0.15 | 0.24 | 0.30 | 0.85 | 0.83 | 0.88 | 0.92 |  |  |  |  |  |
| **FORCE2** | 0.18 | 0.25 | 0.24 | 0.31 | 0.35 | 0.27 | 0.24 | 0.29 | 0.30 | 0.69 | 0.69 | 0.69 | 0.76 | 0.80 |  |  |  |  |
| **COERCE2** | 0.21 | 0.22 | 0.29 | 0.33 | 0.35 | 0.26 | 0.24 | 0.31 | 0.29 | 0.72 | 0.73 | 0.75 | 0.81 | 0.81 | 0.95 |  |  |  |
| **OTHSEX2** | 0.17 | 0.23 | 0.24 | 0.31 | 0.30 | 0.27 | 0.20 | 0.25 | 0.27 | 0.67 | 0.73 | 0.73 | 0.78 | 0.79 | 0.95 | 0.96 |  |  |
| **PORN2** | 0.27 | 0.34 | 0.35 | 0.37 | 0.38 | 0.32 | 0.32 | 0.33 | 0.41 | 0.67 | 0.69 | 0.73 | 0.76 | 0.82 | 0.88 | 0.91 | 0.91 |  |
| **SSCF (N=505)** |  |  |  |  |  |  |  |  |  |  |  |  |  |  |  |  |  |  |
|  | **SLAP0** | **PUSH0** | **HIT0** | **KICK0** | **WEAPON0** | **FORCE0** | **COERCE0** | **OTHSEX0** | **PORN0** | **SLAP2** | **PUSH2** | **HIT2** | **KICK2** | **WEAPON2** | **FORCE2** | **COERCE2** | **OTHSEX2** | **PORN2** |
| **SLAP0** |  |  |  |  |  |  |  |  |  |  |  |  |  |  |  |  |  |  |
| **PUSH0** | 0.72 |  |  |  |  |  |  |  |  |  |  |  |  |  |  |  |  |  |
| **HIT0** | 0.72 | 0.74 |  |  |  |  |  |  |  |  |  |  |  |  |  |  |  |  |
| **KICK0** | 0.67 | 0.75 | 0.80 |  |  |  |  |  |  |  |  |  |  |  |  |  |  |  |
| **WEAPON0** | 0.57 | 0.69 | 0.78 | 0.88 |  |  |  |  |  |  |  |  |  |  |  |  |  |  |
| **FORCE0** | 0.41 | 0.49 | 0.52 | 0.61 | 0.71 |  |  |  |  |  |  |  |  |  |  |  |  |  |
| **COERCE0** | 0.43 | 0.58 | 0.58 | 0.64 | 0.77 | 0.85 |  |  |  |  |  |  |  |  |  |  |  |  |
| **OTHSEX0** | 0.41 | 0.56 | 0.59 | 0.70 | 0.74 | 0.79 | 0.77 |  |  |  |  |  |  |  |  |  |  |  |
| **PORN0** | 0.43 | 0.59 | 0.58 | 0.62 | 0.75 | 0.57 | 0.71 | 0.70 |  |  |  |  |  |  |  |  |  |  |
| **SLAP2** | 0.26 | 0.22 | 0.20 | 0.27 | 0.28 | 0.26 | 0.24 | 0.24 | 0.35 |  |  |  |  |  |  |  |  |  |
| **PUSH2** | 0.35 | 0.35 | 0.27 | 0.31 | 0.33 | 0.22 | 0.28 | 0.24 | 0.24 | 0.74 |  |  |  |  |  |  |  |  |
| **HIT2** | 0.29 | 0.36 | 0.26 | 0.37 | 0.36 | 0.18 | 0.30 | 0.28 | 0.31 | 0.76 | 0.78 |  |  |  |  |  |  |  |
| **KICK2** | 0.38 | 0.38 | 0.29 | 0.37 | 0.40 | 0.31 | 0.32 | 0.31 | 0.33 | 0.75 | 0.80 | 0.85 |  |  |  |  |  |  |
| **WEAPON2** | 0.30 | 0.29 | 0.25 | 0.40 | 0.37 | 0.36 | 0.35 | 0.36 | 0.47 | 0.65 | 0.66 | 0.80 | 0.81 |  |  |  |  |  |
| **FORCE2** | 0.34 | 0.25 | 0.30 | 0.40 | 0.39 | 0.45 | 0.44 | 0.46 | 0.43 | 0.57 | 0.60 | 0.63 | 0.58 | 0.66 |  |  |  |  |
| **COERCE2** | 0.37 | 0.26 | 0.27 | 0.34 | 0.41 | 0.43 | 0.47 | 0.41 | 0.45 | 0.63 | 0.58 | 0.72 | 0.69 | 0.76 | 0.85 |  |  |  |
| **OTHSEX2** | 0.24 | 0.27 | 0.23 | 0.26 | 0.32 | 0.38 | 0.41 | 0.39 | 0.38 | 0.52 | 0.47 | 0.64 | 0.58 | 0.73 | 0.82 | 0.89 |  |  |
| **PORN2** | 0.28 | 0.23 | 0.21 | 0.26 | 0.25 | 0.26 | 0.33 | 0.24 | 0.42 | 0.66 | 0.61 | 0.63 | 0.69 | 0.76 | 0.74 | 0.79 | 0.84 |  |
| **Notes**: IND: Indashyikirwa; SSFC: Stepping Stones and Creating Futures; OMC: One Man Can; RMSEA: Root Mean Square Error of Approximation; CI: Confidence Interval; CFI: Comparative Fit Index; TLI: Tucker Lewis Index. | | | | | | | | | | | | | | | | | | |

**Online Supplement Table 2. Item-Level Correlation by Time and Study Arm.**

| **IND Tx (N=763)** | **SLAP0** | **PUSH0** | **HIT0** | **KICK0** | **WEAPON0** | **FORCE0** | **COERCE0** | **OTHSEX0** | **SLAP2** | **PUSH2** | **HIT2** | **KICK2** | **WEAPON2** | **FORCE2** | **COERCE2** | **OTHSEX2** |  |  |
| --- | --- | --- | --- | --- | --- | --- | --- | --- | --- | --- | --- | --- | --- | --- | --- | --- | --- | --- |
| SLAP0 |  |  |  |  |  |  |  |  |  |  |  |  |  |  |  |  |  |  |
| PUSH0 | 0.77 |  |  |  |  |  |  |  |  |  |  |  |  |  |  |  |  |  |
| HIT0 | 0.78 | 0.76 |  |  |  |  |  |  |  |  |  |  |  |  |  |  |  |  |
| KICK0 | 0.57 | 0.50 | 0.69 |  |  |  |  |  |  |  |  |  |  |  |  |  |  |  |
| WEAPON0 | 0.18 | 0.38 | 0.30 | 0.38 |  |  |  |  |  |  |  |  |  |  |  |  |  |  |
| FORCE0 | 0.44 | 0.45 | 0.40 | 0.38 | 0.48 |  |  |  |  |  |  |  |  |  |  |  |  |  |
| COERCE0 | 0.41 | 0.43 | 0.34 | 0.24 | 0.14 | 0.82 |  |  |  |  |  |  |  |  |  |  |  |  |
| OTHSEX0 | 0.18 | 0.28 | 0.15 | 0.18 | 0.19 | 0.14 | 0.32 |  |  |  |  |  |  |  |  |  |  |  |
| SLAP2 | 0.36 | 0.36 | 0.29 | 0.33 | 0.01 | 0.30 | 0.17 | 0.10 |  |  |  |  |  |  |  |  |  |  |
| PUSH2 | 0.50 | 0.48 | 0.34 | 0.30 | 0.25 | 0.27 | 0.21 | 0.10 | 0.73 |  |  |  |  |  |  |  |  |  |
| HIT2 | 0.30 | 0.31 | 0.35 | 0.03 | 0.06 | 0.31 | 0.10 | 0.01 | 0.82 | 0.73 |  |  |  |  |  |  |  |  |
| KICK2 | 0.15 | 0.17 | 0.27 | 0.45 | 0.27 | 0.06 | -0.17 | 0.43 | 0.66 | 0.83 | 0.77 |  |  |  |  |  |  |  |
| WEAPON2 | 0.20 | 0.00 | -0.12 | 0.15 | 0.30 | 0.11 | -0.25 | 0.13 | 0.58 | 0.72 | 0.71 | 0.84 |  |  |  |  |  |  |
| FORCE2 | 0.26 | 0.16 | 0.22 | 0.17 | 0.04 | 0.46 | 0.34 | -0.02 | 0.50 | 0.57 | 0.66 | 0.75 | 0.69 |  |  |  |  |  |
| COERCE2 | 0.30 | 0.22 | 0.14 | 0.28 | -0.04 | 0.40 | 0.41 | -0.10 | 0.41 | 0.49 | 0.55 | 0.68 | 0.44 | 0.90 |  |  |  |  |
| OTHSEX2 | 0.09 | 0.14 | 0.22 | 0.02 | 0.35 | 0.20 | 0.12 | 0.32 | 0.37 | 0.25 | 0.56 | 0.64 | 0.68 | 0.48 | 0.38 |  |  |  |
| **IND Ctl (N=774)** | **SLAP0** | **PUSH0** | **HIT0** | **KICK0** | **WEAPON0** | **FORCE0** | **COERCE0** | **OTHSEX0** | **SLAP2** | **PUSH2** | **HIT2** | **KICK2** | **WEAPON2** | **FORCE2** | **COERCE2** | **OTHSEX2** |  |  |
| SLAP0 |  |  |  |  |  |  |  |  |  |  |  |  |  |  |  |  |  |  |
| PUSH0 | 0.70 |  |  |  |  |  |  |  |  |  |  |  |  |  |  |  |  |  |
| HIT0 | 0.84 | 0.71 |  |  |  |  |  |  |  |  |  |  |  |  |  |  |  |  |
| KICK0 | 0.66 | 0.76 | 0.84 |  |  |  |  |  |  |  |  |  |  |  |  |  |  |  |
| WEAPON0 | 0.72 | 0.66 | 0.84 | 0.88 |  |  |  |  |  |  |  |  |  |  |  |  |  |  |
| FORCE0 | 0.56 | 0.52 | 0.64 | 0.70 | 0.79 |  |  |  |  |  |  |  |  |  |  |  |  |  |
| COERCE0 | 0.52 | 0.51 | 0.55 | 0.66 | 0.66 | 0.86 |  |  |  |  |  |  |  |  |  |  |  |  |
| OTHSEX0 | 0.30 | 0.15 | 0.41 | 0.57 | 0.64 | 0.50 | 0.53 |  |  |  |  |  |  |  |  |  |  |  |
| SLAP2 | 0.60 | 0.41 | 0.48 | 0.53 | 0.55 | 0.45 | 0.36 | 0.32 |  |  |  |  |  |  |  |  |  |  |
| PUSH2 | 0.57 | 0.51 | 0.56 | 0.53 | 0.71 | 0.42 | 0.45 | 0.27 | 0.73 |  |  |  |  |  |  |  |  |  |
| HIT2 | 0.57 | 0.52 | 0.55 | 0.61 | 0.66 | 0.65 | 0.61 | 0.37 | 0.79 | 0.75 |  |  |  |  |  |  |  |  |
| KICK2 | 0.60 | 0.44 | 0.75 | 0.75 | 0.89 | 0.52 | 0.44 | 0.45 | 0.77 | 0.82 | 0.85 |  |  |  |  |  |  |  |
| WEAPON2 | 0.38 | 0.17 | 0.33 | 0.50 | 0.41 | 0.06 | -0.02 | 0.15 | 0.61 | 0.36 | 0.47 | 0.80 |  |  |  |  |  |  |
| FORCE2 | 0.32 | 0.33 | 0.45 | 0.41 | 0.59 | 0.66 | 0.57 | 0.42 | 0.52 | 0.56 | 0.66 | 0.76 | 0.48 |  |  |  |  |  |
| COERCE2 | 0.33 | 0.18 | 0.37 | 0.49 | 0.39 | 0.54 | 0.53 | 0.37 | 0.52 | 0.43 | 0.61 | 0.66 | 0.48 | 0.85 |  |  |  |  |
| OTHSEX2 | 0.32 | 0.09 | 0.28 | 0.10 | 0.22 | 0.26 | 0.17 | 0.28 | 0.51 | 0.29 | 0.34 | 0.49 | 0.68 | 0.55 | 0.51 |  |  |  |
| **OMC Tx (N=746)** | **SLAP0** | **PUSH0** | **HIT0** | **KICK0** | **WEAPON0** | **FORCE0** | **COERCE0** | **OTHSEX0** | **PORN0** | **SLAP2** | **PUSH2** | **HIT2** | **KICK2** | **WEAPON2** | **FORCE2** | **COERCE2** | **OTHSEX2** | **PORN2** |
| SLAP0 |  |  |  |  |  |  |  |  |  |  |  |  |  |  |  |  |  |  |
| PUSH0 | 0.76 |  |  |  |  |  |  |  |  |  |  |  |  |  |  |  |  |  |
| HIT0 | 0.77 | 0.79 |  |  |  |  |  |  |  |  |  |  |  |  |  |  |  |  |
| KICK0 | 0.72 | 0.75 | 0.79 |  |  |  |  |  |  |  |  |  |  |  |  |  |  |  |
| WEAPON0 | 0.61 | 0.67 | 0.73 | 0.78 |  |  |  |  |  |  |  |  |  |  |  |  |  |  |
| FORCE0 | 0.51 | 0.54 | 0.61 | 0.58 | 0.61 |  |  |  |  |  |  |  |  |  |  |  |  |  |
| COERCE0 | 0.55 | 0.57 | 0.65 | 0.61 | 0.67 | 0.85 |  |  |  |  |  |  |  |  |  |  |  |  |
| OTHSEX0 | 0.57 | 0.60 | 0.65 | 0.68 | 0.68 | 0.77 | 0.84 |  |  |  |  |  |  |  |  |  |  |  |
| PORN0 | 0.48 | 0.60 | 0.65 | 0.66 | 0.67 | 0.67 | 0.76 | 0.84 |  |  |  |  |  |  |  |  |  |  |
| SLAP2 | 0.26 | 0.30 | 0.32 | 0.33 | 0.19 | 0.33 | 0.24 | 0.29 | 0.28 |  |  |  |  |  |  |  |  |  |
| PUSH2 | 0.29 | 0.38 | 0.36 | 0.32 | 0.26 | 0.26 | 0.32 | 0.30 | 0.35 | 0.88 |  |  |  |  |  |  |  |  |
| HIT2 | 0.28 | 0.33 | 0.41 | 0.37 | 0.19 | 0.27 | 0.23 | 0.24 | 0.27 | 0.92 | 0.86 |  |  |  |  |  |  |  |
| KICK2 | 0.20 | 0.30 | 0.34 | 0.39 | 0.29 | 0.27 | 0.18 | 0.28 | 0.30 | 0.90 | 0.83 | 0.92 |  |  |  |  |  |  |
| WEAPON2 | 0.27 | 0.39 | 0.42 | 0.37 | 0.35 | 0.26 | 0.19 | 0.28 | 0.36 | 0.83 | 0.84 | 0.87 | 0.91 |  |  |  |  |  |
| FORCE2 | 0.22 | 0.36 | 0.30 | 0.32 | 0.41 | 0.33 | 0.34 | 0.34 | 0.38 | 0.67 | 0.70 | 0.69 | 0.78 | 0.84 |  |  |  |  |
| COERCE2 | 0.24 | 0.28 | 0.33 | 0.36 | 0.45 | 0.33 | 0.28 | 0.33 | 0.31 | 0.72 | 0.74 | 0.77 | 0.82 | 0.83 | 0.95 |  |  |  |
| OTHSEX2 | 0.15 | 0.29 | 0.26 | 0.30 | 0.39 | 0.36 | 0.29 | 0.32 | 0.29 | 0.70 | 0.74 | 0.74 | 0.81 | 0.85 | 0.95 | 0.97 |  |  |
| PORN2 | 0.24 | 0.37 | 0.39 | 0.42 | 0.42 | 0.38 | 0.38 | 0.41 | 0.46 | 0.68 | 0.72 | 0.75 | 0.78 | 0.84 | 0.88 | 0.92 | 0.93 |  |
| **OMC Tx (N=714)** | **SLAP0** | **PUSH0** | **HIT0** | **KICK0** | **WEAPON0** | **FORCE0** | **COERCE0** | **OTHSEX0** | **PORN0** | **SLAP2** | **PUSH2** | **HIT2** | **KICK2** | **WEAPON2** | **FORCE2** | **COERCE2** | **OTHSEX2** | **PORN2** |
| SLAP0 |  |  |  |  |  |  |  |  |  |  |  |  |  |  |  |  |  |  |
| PUSH0 | 0.72 |  |  |  |  |  |  |  |  |  |  |  |  |  |  |  |  |  |
| HIT0 | 0.75 | 0.67 |  |  |  |  |  |  |  |  |  |  |  |  |  |  |  |  |
| KICK0 | 0.70 | 0.73 | 0.80 |  |  |  |  |  |  |  |  |  |  |  |  |  |  |  |
| WEAPON0 | 0.68 | 0.67 | 0.76 | 0.82 |  |  |  |  |  |  |  |  |  |  |  |  |  |  |
| FORCE0 | 0.48 | 0.56 | 0.54 | 0.56 | 0.53 |  |  |  |  |  |  |  |  |  |  |  |  |  |
| COERCE0 | 0.59 | 0.58 | 0.64 | 0.66 | 0.63 | 0.76 |  |  |  |  |  |  |  |  |  |  |  |  |
| OTHSEX0 | 0.61 | 0.59 | 0.72 | 0.68 | 0.68 | 0.74 | 0.76 |  |  |  |  |  |  |  |  |  |  |  |
| PORN0 | 0.44 | 0.50 | 0.56 | 0.62 | 0.63 | 0.66 | 0.63 | 0.68 |  |  |  |  |  |  |  |  |  |  |
| SLAP2 | 0.34 | 0.28 | 0.21 | 0.37 | 0.31 | 0.28 | 0.13 | 0.21 | 0.26 |  |  |  |  |  |  |  |  |  |
| PUSH2 | 0.31 | 0.30 | 0.30 | 0.33 | 0.28 | 0.31 | 0.14 | 0.27 | 0.29 | 0.91 |  |  |  |  |  |  |  |  |
| HIT2 | 0.29 | 0.30 | 0.32 | 0.42 | 0.35 | 0.30 | 0.21 | 0.28 | 0.34 | 0.93 | 0.86 |  |  |  |  |  |  |  |
| KICK2 | 0.19 | 0.23 | 0.25 | 0.39 | 0.23 | 0.24 | 0.14 | 0.19 | 0.33 | 0.88 | 0.85 | 0.91 |  |  |  |  |  |  |
| WEAPON2 | 0.16 | 0.31 | 0.27 | 0.39 | 0.23 | 0.22 | 0.12 | 0.21 | 0.27 | 0.87 | 0.83 | 0.90 | 0.94 |  |  |  |  |  |
| FORCE2 | 0.14 | 0.11 | 0.17 | 0.31 | 0.28 | 0.20 | 0.13 | 0.23 | 0.21 | 0.71 | 0.67 | 0.69 | 0.74 | 0.77 |  |  |  |  |
| COERCE2 | 0.17 | 0.16 | 0.24 | 0.29 | 0.21 | 0.16 | 0.19 | 0.30 | 0.27 | 0.72 | 0.73 | 0.74 | 0.81 | 0.81 | 0.96 |  |  |  |
| OTHSEX2 | 0.19 | 0.17 | 0.22 | 0.31 | 0.18 | 0.17 | 0.09 | 0.17 | 0.25 | 0.64 | 0.72 | 0.72 | 0.74 | 0.74 | 0.94 | 0.96 |  |  |
| PORN2 | 0.31 | 0.31 | 0.32 | 0.33 | 0.33 | 0.25 | 0.26 | 0.24 | 0.36 | 0.66 | 0.66 | 0.71 | 0.74 | 0.80 | 0.87 | 0.90 | 0.89 |  |
| **SSCF Tx (N=237)** | **SLAP0** | **PUSH0** | **HIT0** | **KICK0** | **WEAPON0** | **FORCE0** | **COERCE0** | **OTHSEX0** | **PORN0** | **SLAP2** | **PUSH2** | **HIT2** | **KICK2** | **WEAPON2** | **FORCE2** | **COERCE2** | **OTHSEX2** | **PORN2** |
| SLAP0 |  |  |  |  |  |  |  |  |  |  |  |  |  |  |  |  |  |  |
| PUSH0 | 0.80 |  |  |  |  |  |  |  |  |  |  |  |  |  |  |  |  |  |
| HIT0 | 0.63 | 0.73 |  |  |  |  |  |  |  |  |  |  |  |  |  |  |  |  |
| KICK0 | 0.55 | 0.73 | 0.82 |  |  |  |  |  |  |  |  |  |  |  |  |  |  |  |
| WEAPON0 | 0.54 | 0.72 | 0.83 | 0.89 |  |  |  |  |  |  |  |  |  |  |  |  |  |  |
| FORCE0 | 0.41 | 0.58 | 0.59 | 0.74 | 0.78 |  |  |  |  |  |  |  |  |  |  |  |  |  |
| COERCE0 | 0.43 | 0.56 | 0.55 | 0.68 | 0.80 | 0.88 |  |  |  |  |  |  |  |  |  |  |  |  |
| OTHSEX0 | 0.41 | 0.65 | 0.66 | 0.79 | 0.78 | 0.85 | 0.83 |  |  |  |  |  |  |  |  |  |  |  |
| PORN0 | 0.43 | 0.72 | 0.68 | 0.70 | 0.85 | 0.77 | 0.84 | 0.80 |  |  |  |  |  |  |  |  |  |  |
| SLAP2 | 0.18 | 0.25 | 0.19 | 0.31 | 0.22 | 0.28 | 0.25 | 0.28 | 0.27 |  |  |  |  |  |  |  |  |  |
| PUSH2 | 0.30 | 0.34 | 0.29 | 0.34 | 0.31 | 0.23 | 0.33 | 0.32 | 0.31 | 0.82 |  |  |  |  |  |  |  |  |
| HIT2 | 0.35 | 0.45 | 0.35 | 0.41 | 0.36 | 0.29 | 0.44 | 0.38 | 0.45 | 0.77 | 0.81 |  |  |  |  |  |  |  |
| KICK2 | 0.39 | 0.46 | 0.33 | 0.44 | 0.40 | 0.34 | 0.43 | 0.44 | 0.41 | 0.79 | 0.84 | 0.88 |  |  |  |  |  |  |
| WEAPON2 | 0.34 | 0.32 | 0.35 | 0.50 | 0.45 | 0.40 | 0.39 | 0.50 | 0.51 | 0.68 | 0.77 | 0.86 | 0.90 |  |  |  |  |  |
| FORCE2 | 0.36 | 0.27 | 0.30 | 0.46 | 0.51 | 0.50 | 0.53 | 0.59 | 0.56 | 0.51 | 0.68 | 0.70 | 0.69 | 0.74 |  |  |  |  |
| COERCE2 | 0.45 | 0.48 | 0.36 | 0.50 | 0.64 | 0.59 | 0.66 | 0.67 | 0.72 | 0.58 | 0.70 | 0.77 | 0.77 | 0.81 | 0.87 |  |  |  |
| OTHSEX2 | 0.32 | 0.27 | 0.39 | 0.31 | 0.56 | 0.36 | 0.49 | 0.45 | 0.60 | 0.47 | 0.58 | 0.70 | 0.66 | 0.81 | 0.85 | 0.91 |  |  |
| PORN2 | 0.21 | 0.24 | 0.27 | 0.27 | 0.42 | 0.37 | 0.36 | 0.26 | 0.53 | 0.57 | 0.70 | 0.65 | 0.80 | 0.79 | 0.78 | 0.79 | 0.84 |  |
| **SSCF Ctl (N=268)** | **SLAP0** | **PUSH0** | **HIT0** | **KICK0** | **WEAPON0** | **FORCE0** | **COERCE0** | **OTHSEX0** | **PORN0** | **SLAP2** | **PUSH2** | **HIT2** | **KICK2** | **WEAPON2** | **FORCE2** | **COERCE2** | **OTHSEX2** | **PORN2** |
| SLAP0 |  |  |  |  |  |  |  |  |  |  |  |  |  |  |  |  |  |  |
| PUSH0 | 0.64 |  |  |  |  |  |  |  |  |  |  |  |  |  |  |  |  |  |
| HIT0 | 0.80 | 0.75 |  |  |  |  |  |  |  |  |  |  |  |  |  |  |  |  |
| KICK0 | 0.77 | 0.77 | 0.78 |  |  |  |  |  |  |  |  |  |  |  |  |  |  |  |
| WEAPON0 | 0.60 | 0.66 | 0.72 | 0.88 |  |  |  |  |  |  |  |  |  |  |  |  |  |  |
| FORCE0 | 0.42 | 0.41 | 0.47 | 0.49 | 0.65 |  |  |  |  |  |  |  |  |  |  |  |  |  |
| COERCE0 | 0.43 | 0.61 | 0.60 | 0.59 | 0.74 | 0.83 |  |  |  |  |  |  |  |  |  |  |  |  |
| OTHSEX0 | 0.41 | 0.49 | 0.54 | 0.61 | 0.72 | 0.74 | 0.72 |  |  |  |  |  |  |  |  |  |  |  |
| PORN0 | 0.44 | 0.47 | 0.49 | 0.53 | 0.61 | 0.34 | 0.57 | 0.60 |  |  |  |  |  |  |  |  |  |  |
| SLAP2 | 0.33 | 0.19 | 0.22 | 0.23 | 0.34 | 0.23 | 0.23 | 0.21 | 0.41 |  |  |  |  |  |  |  |  |  |
| PUSH2 | 0.39 | 0.37 | 0.26 | 0.28 | 0.35 | 0.20 | 0.24 | 0.18 | 0.18 | 0.67 |  |  |  |  |  |  |  |  |
| HIT2 | 0.24 | 0.28 | 0.19 | 0.33 | 0.36 | 0.09 | 0.17 | 0.20 | 0.16 | 0.75 | 0.76 |  |  |  |  |  |  |  |
| KICK2 | 0.39 | 0.31 | 0.26 | 0.32 | 0.42 | 0.28 | 0.23 | 0.19 | 0.27 | 0.71 | 0.77 | 0.82 |  |  |  |  |  |  |
| WEAPON2 | 0.25 | 0.26 | 0.14 | 0.31 | 0.29 | 0.33 | 0.31 | 0.24 | 0.43 | 0.63 | 0.54 | 0.74 | 0.73 |  |  |  |  |  |
| FORCE2 | 0.32 | 0.24 | 0.31 | 0.35 | 0.27 | 0.40 | 0.35 | 0.35 | 0.29 | 0.61 | 0.53 | 0.56 | 0.48 | 0.58 |  |  |  |  |
| COERCE2 | 0.31 | 0.08 | 0.21 | 0.19 | 0.16 | 0.30 | 0.28 | 0.16 | 0.16 | 0.66 | 0.48 | 0.68 | 0.62 | 0.71 | 0.83 |  |  |  |
| OTHSEX2 | 0.17 | 0.27 | 0.09 | 0.22 | 0.07 | 0.39 | 0.35 | 0.34 | 0.16 | 0.54 | 0.38 | 0.60 | 0.51 | 0.66 | 0.80 | 0.87 |  |  |
| PORN2 | 0.34 | 0.23 | 0.17 | 0.25 | 0.06 | 0.17 | 0.30 | 0.22 | 0.33 | 0.72 | 0.53 | 0.62 | 0.59 | 0.74 | 0.70 | 0.79 | 0.84 |  |
| **Notes**: IND: Indashyikirwa; SSFC: Stepping Stones and Creating Futures; OMC: One Man Can; RMSEA: Root Mean Square Error of Approximation; CI: Confidence Interval; CFI: Comparative Fit Index; TLI: Tucker Lewis Index; Tx: Treatment Arm; Ctl: Control Arm. | | | | | | | | | | | | | | | | | | |

**Table 3. Average Item Correlation Within and Across IPV Types, by Study, Study Arm, and Time.**

|  | **IND (N=1537)** | | | | | | | | | | | |
| --- | --- | --- | --- | --- | --- | --- | --- | --- | --- | --- | --- | --- |
|  | **Mean (N=1537)** | | | | **Tx (N=763)** | | | | **Ctl (N=774)** | | | |
|  | **Baseline** | | **Endline** | | **Baseline** | | **Endline** | | **Baseline** | | **Endline** | |
|  | **Within** | **Across** | **Within** | **Across** | **Within** | **Across** | **Within** | **Across** | **Within** | **Across** | **Within** | **Across** |
| SLAP | 0.68 | 0.40 | 0.71 | 0.47 | 0.57 | 0.34 | 0.69 | 0.43 | 0.73 | 0.46 | 0.72 | 0.52 |
| PUSH | 0.67 | 0.39 | 0.72 | 0.43 | 0.60 | 0.39 | 0.75 | 0.43 | 0.71 | 0.39 | 0.66 | 0.43 |
| HIT | 0.75 | 0.42 | 0.74 | 0.56 | 0.63 | 0.30 | 0.76 | 0.59 | 0.81 | 0.53 | 0.71 | 0.54 |
| KICK | 0.70 | 0.51 | 0.80 | 0.66 | 0.53 | 0.27 | 0.77 | 0.69 | 0.79 | 0.64 | 0.81 | 0.64 |
| WEAPON | 0.63 | 0.55 | 0.65 | 0.57 | 0.31 | 0.27 | 0.71 | 0.60 | 0.78 | 0.70 | 0.56 | 0.55 |
| FORCE | 0.59 | 0.55 | 0.69 | 0.61 | 0.48 | 0.43 | 0.69 | 0.63 | 0.68 | 0.64 | 0.70 | 0.59 |
| COERCE | 0.64 | 0.47 | 0.66 | 0.52 | 0.57 | 0.31 | 0.64 | 0.51 | 0.69 | 0.58 | 0.68 | 0.54 |
| OTHSEX | 0.39 | 0.34 | 0.48 | 0.49 | 0.23 | 0.19 | 0.43 | 0.50 | 0.52 | 0.41 | 0.53 | 0.46 |
| PORN | NA | NA | NA | NA | NA | NA | NA | NA | NA | NA | NA | NA |
| **SSCF (N=505)** | | | | | | | | | | | | |
|  | **Mean (N=505)** | | | | **Tx (N=237)** | | | | **Ctl (N=268)** | | | |
|  | **Baseline** | | **Endline** | | **Baseline** | | **Endline** | | **Baseline** | | **Endline** | |
|  | **Within** | **Across** | **Within** | **Across** | **Within** | **Across** | **Within** | **Across** | **Within** | **Across** | **Within** | **Across** |
| SLAP | 0.67 | 0.42 | 0.73 | 0.59 | 0.63 | 0.42 | 0.77 | 0.53 | 0.70 | 0.42 | 0.69 | 0.63 |
| PUSH | 0.72 | 0.56 | 0.75 | 0.57 | 0.74 | 0.63 | 0.81 | 0.66 | 0.70 | 0.49 | 0.68 | 0.48 |
| HIT | 0.76 | 0.57 | 0.80 | 0.65 | 0.75 | 0.62 | 0.83 | 0.71 | 0.76 | 0.52 | 0.77 | 0.61 |
| KICK | 0.78 | 0.64 | 0.80 | 0.64 | 0.75 | 0.73 | **0.85** | 0.73 | 0.80 | 0.56 | 0.76 | 0.55 |
| WEAPON | 0.73 | 0.74 | 0.73 | 0.73 | 0.74 | 0.80 | 0.80 | 0.79 | 0.72 | 0.68 | 0.66 | 0.67 |
| FORCE | 0.74 | 0.55 | 0.80 | 0.61 | 0.83 | 0.62 | 0.83 | 0.67 | 0.63 | 0.49 | 0.78 | 0.55 |
| COERCE | 0.78 | 0.60 | 0.84 | 0.67 | 0.85 | 0.60 | **0.86** | 0.73 | 0.71 | 0.59 | 0.83 | 0.63 |
| OTHSEX | 0.75 | 0.60 | 0.85 | 0.59 | 0.83 | 0.66 | **0.87** | 0.64 | 0.69 | 0.55 | 0.83 | 0.54 |
| PORN | 0.66 | 0.59 | 0.79 | 0.67 | 0.81 | 0.68 | 0.80 | 0.70 | 0.50 | 0.51 | 0.78 | 0.64 |
| **OMC (N=1460)** | | | | | | | | | | | | |
|  | **Mean (N=1460)** | | | | **Tx (N=746)** | | | | **Ctl (N=714)** | | | |
|  | **Baseline** | | **Endline** | | **Baseline** | | **Endline** | | **Baseline** | | **Endline** | |
|  | **Within** | **Across** | **Within** | **Across** | **Within** | **Across** | **Within** | **Across** | **Within** | **Across** | **Within** | **Across** |
| SLAP | 0.71 | 0.53 | **0.89** | 0.69 | 0.71 | 0.53 | **0.88** | 0.69 | 0.71 | 0.53 | **0.90** | 0.68 |
| PUSH | 0.72 | 0.57 | **0.86** | 0.71 | 0.74 | 0.58 | **0.85** | 0.72 | 0.70 | 0.55 | **0.86** | 0.70 |
| HIT | 0.76 | 0.63 | **0.90** | 0.73 | 0.77 | 0.64 | **0.89** | 0.74 | 0.74 | 0.62 | **0.90** | 0.71 |
| KICK | 0.76 | 0.63 | **0.89** | 0.78 | 0.76 | 0.63 | **0.89** | 0.80 | 0.76 | 0.63 | **0.89** | 0.76 |
| WEAPON | 0.72 | 0.64 | **0.87** | 0.81 | 0.70 | 0.66 | **0.86** | 0.84 | 0.73 | 0.62 | **0.89** | 0.78 |
| FORCE | 0.74 | 0.55 | **0.92** | 0.73 | 0.76 | 0.57 | **0.93** | 0.74 | 0.72 | 0.53 | **0.92** | 0.72 |
| COERCE | 0.77 | 0.61 | **0.94** | 0.77 | 0.82 | 0.61 | **0.94** | 0.77 | 0.71 | 0.62 | **0.94** | 0.76 |
| OTHSEX | 0.78 | 0.65 | **0.94** | 0.74 | 0.82 | 0.64 | **0.95** | 0.77 | 0.73 | 0.66 | **0.93** | 0.71 |
| PORN | 0.71 | 0.58 | **0.90** | 0.73 | 0.75 | 0.61 | **0.91** | 0.75 | 0.66 | 0.55 | **0.89** | 0.71 |
| Notes: IND: Indashyikirwa; SSFC: Stepping Stones and Creating Futures; OMC: One Man Can; Tx: Treatment; Ctl: Control; Bold items reflect correlations ≥ 0.85. | | | | | | | | | | | | |

**Online Supplement Table 4. Item-Level Exploratory Factor Analysis Results by Study, Baseline.**

| **IND** | **One Factor** | **Two-Factor** | |
| --- | --- | --- | --- |
| N | 768 | | |
| CHI SQUARE | 98.34 | 16.86 | |
| DEGEES OF FREEDOM | 20 | 13 | |
| P-VALUE | 0 | 0.21 | |
| RMSEA | 0.07 | 0.02 | |
| RMSEA 90%CI | (0.06, 0.09) | (0.00, 0.04) | |
| CFI | 0.94 | 1.00 | |
| TLI | 0.92 | 0.99 | |
| **ITEM** | **F1** | **F1** | **F2** |
| SLAP0 | 0.84 | 0.98 | -0.16 |
| PUSH0 | 0.79 | 0.79 | 0.06 |
| HIT0 | 0.90 | 0.92 | 0.01 |
| KICK0 | 0.85 | 0.63 | 0.35 |
| WEAPON0 | 0.83 | 0.33 | 0.63 |
| FORCE0 | 0.87 | 0.11 | 0.87 |
| COERCE0 | 0.83 | -0.00 | 0.90 |
| OTHSEX0 | 0.40 | -0.17 | 0.63 |
| **OMC** | **One Factor** | **Two-Factor** | |
| N | 728 | | |
| CHI SQUARE | 68.20 | 26.96 | |
| DEGEES OF FREEDOM | 27 | 19 | |
| P-VALUE | 0.00 | 0.11 | |
| RMSEA | 0.05 | 0.02 | |
| RMSEA 90%CI | (0.03, 0.06) | (0.00, 0.04) | |
| CFI | 0.99 | 1.00 |  |
| TLI | 0.98 | 1.00 |  |
| **ITEM** | **F1** | **F1** | **F2** |
| SLAP0 | 0.75 | 0.81 | -0.01 |
| PUSH0 | 0.79 | 0.85 | -0.01 |
| HIT0 | 0.86 | 0.65 | 0.28 |
| KICK0 | 0.91 | 0.78 | 0.18 |
| WEAPON0 | 0.83 | 0.61 | 0.28 |
| FORCE0 | 0.82 | -0.07 | 0.95 |
| COERCE0 | 0.86 | 0.01 | 0.89 |
| OTHSEX0 | 0.87 | 0.25 | 0.70 |
| PORN0 | 0.76 | 0.28 | 0.54 |
| **SSCF** | **One Factor** | **Two-Factor** | |
| N | 253 | | |
| CHI SQUARE | 117.06 | 25.15 | |
| DEGEES OF FREEDOM | 27 | 19 | |
| P-VALUE | 0.00 | 0.16 | |
| RMSEA | 0.12 | 0.04 | |
| RMSEA 90%CI | (0.09, 0.14) | (0.000, 0.070) | |
| CFI | 0.95 | 1.00 | |
| TLI | 0.94 | 0.99 | |
| **ITEM** | **F1** | **F1** | **F2** |
| SLAP0 | 0.72 | 1.00 | -0.28 |
| PUSH0 | 0.84 | 0.86 | 0.00 |
| HIT0 | 0.81 | 0.87 | 0.01 |
| KICK0 | 0.88 | 0.82 | 0.16 |
| WEAPON0 | 0.92 | 0.55 | 0.51 |
| FORCE0 | 0.86 | -0.01 | 0.98 |
| COERCE0 | 0.88 | 0.17 | 0.79 |
| OTHSEX0 | 0.79 | 0.31 | 0.6 |
| PORN0 | 0.78 | 0.45 | 0.46 |
| Notes: IND: Indashyikirwa; SSFC: Stepping Stones and Creating Futures; OMC: One Man Can; RMSEA: Root Mean Square Error of Approximation; CI: Confidence Interval; CFI: Comparative Fit Index; TLI: Tucker Lewis Index; F1: Factor 1; F2: Factor 2. | | | |

**Online Supplement Table 5. Item-Level Confirmatory Factor Analysis Results by Study, Baseline.**

| **IND** | **Two-Factor** | |
| --- | --- | --- |
| N |  | |
| CHI SQUARE | 29.56 | |
| DEGEES OF FREEDOM | 19 | |
| P-VALUE | 0.06 | |
| RMSEA | 0.03 | |
| RMSEA 90%CI | (0.00, 0.05) | |
| CFI | 0.99 | |
| TLI | 0.98 | |
| **ITEM** | **F1** | **F2** |
| SLAP0 | 0.89 |  |
| PUSH0 | 0.82 |  |
| HIT0 | 0.89 |  |
| KICK0 | 0.75 |  |
| WEAPON0 | 0.76 |  |
| FORCE0 |  | 0.91 |
| COERCE0 |  | 0.90 |
| OTHSEX0 |  | 0.52 |
| **SSCF** | **Two-Factor** | |
| N | 251 | |
| CHI SQUARE | 24.76 | |
| DEGEES OF FREEDOM | 26 | |
| P-VALUE | 0.53 | |
| RMSEA | 0.00 | |
| RMSEA 90%CI | (0.00, 0.05) | |
| CFI | 1.00 | |
| TLI | 1.00 | |
| **ITEM** | **F1** | **F2** |
| SLAP0 | 0.744 |  |
| PUSH0 | 0.821 |  |
| HIT0 | 0.869 |  |
| KICK0 | 0.924 |  |
| WEAPON0 | 1.001 |  |
| FORCE0 |  | 0.869 |
| COERCE0 |  | 0.935 |
| OTHSEX0 |  | 0.923 |
| PORN0 |  | 0.797 |
| **OMC** | **Two-Factor** | |
| N | 731 | |
| CHI SQUARE | 31.13 | |
| DEGEES OF FREEDOM | 26 | |
| P-VALUE | 0.22 | |
| RMSEA | 0.02 | |
| RMSEA 90%CI | (0.00, 0.04) | |
| CFI | 1.00 | |
| TLI | 1.00 | |
| **ITEM** | **F1** | **F2** |
| SLAP0 | 0.82 |  |
| PUSH0 | 0.9 |  |
| HIT0 | 0.92 |  |
| KICK0 | 0.88 |  |
| WEAPON0 | 0.89 |  |
| FORCE0 |  | 0.82 |
| COERCE0 |  | 0.90 |
| OTHSEX0 |  | 0.92 |
| PORN0 |  | 0.86 |
| Notes: IND: Indashyikirwa; SSFC: Stepping Stones and Creating Futures; OMC: One Man Can; RMSEA: Root Mean Square Error of Approximation; CI: Confidence Interval; CFI: Comparative Fit Index; TLI: Tucker Lewis Index; F1: Factor 1; F2: Factor 2. | | |

**Online Supplement Table 6. Measurement Invariance Testing of the Correlated Two-Factor Model by Study.**

| **IND** | | | | | | | | | |
| --- | --- | --- | --- | --- | --- | --- | --- | --- | --- |
| **Model** | **χ2** | **d.f.** | **RMSEA** | **CFI** | **TLI** | **Δχ2 (diff test)** | **ΔRMSEA** | **ΔCFI** | **ΔTLI** |
| Baseline cross-arm (n=1537) |  |  |  |  |  |  |  |  |  |
| Configural | 53.0 | 38 | 0.023 | 0.995 | 0.992 | -- | -- | -- | -- |
| Scalar | 57.4 | 42 | 0.022 | 0.995 | 0.993 | -- | -- | -- | -- |
| Configural v scalar | 5.3 | 4 | -- | -- | -- | 0.258 | -0.001 | 0.000 | 0.001 |
| Endline cross-arm (n=1534) |  |  |  |  |  |  |  |  |  |
| Configural | 70.7 | 38 | 0.033 | 0.984 | 0.977 | -- | -- | -- | -- |
| Scalar | 70.8 | 42 | 0.030 | 0.986 | 0.982 | -- | -- | -- | -- |
| Configural v scalar | 3.1 | 4 | -- | -- | -- | 0.547 | -0.003 | 0.002 | 0.005 |
| Cross-time (B/E) (n=1537) |  |  |  |  |  |  |  |  |  |
| Configural | 161.0 | 98 | 0.020 | 0.981 | 0.977 | -- | -- | -- | -- |
| Scalar | 211.9 | 112 | 0.024 | 0.971 | 0.968 | -- | -- | -- | -- |
| Configural v scalar | 56.0 | 14 | -- | -- | -- | 0.000 | 0.004 | -0.010 | -0.009 |
| Cross-time treatment (B/E) (n=763) |  |  |  |  |  |  |  |  |  |
| Configural | 114.0 | 98 | 0.015 | 0.989 | 0.987 | -- | -- | -- | -- |
| Scalar | 171.9 | 112 | 0.026 | 0.959 | 0.956 | -- | -- | -- | -- |
| Configural v scalar | 63.6 | 14 | -- | -- | -- | 0.000 | 0.011 | -0.030 | -0.031 |
| Cross-time control (B/E) (n=774) |  |  |  |  |  |  |  |  |  |
| Configural | 116.1 | 98 | 0.015 | 0.991 | 0.989 | -- | -- | -- | -- |
| Scalar | 140.9 | 112 | 0.018 | 0.986 | 0.985 | -- | -- | -- | -- |
| Configural v scalar | 27.6 | 14 | -- | -- | -- | 0.016 | 0.003 | -0.005 | -0.004 |
| **SSCF** | | | | | | | | | |
| **Model** | **χ2** | **d.f** | **RMSEA** | **CFI** | **TLI** | **Δχ2 (diff test)** | **ΔRMSEA** | **ΔCFI** | **ΔTLI** |
| Baseline cross-arm (n=504) |  |  |  |  |  |  |  |  |  |
| Configural | 112.3 | 52 | 0.068 | 0.987 | 0.982 | -- | -- | -- | -- |
| Scalar | 114.4 | 57 | 0.063 | 0.988 | 0.985 | -- | -- | -- | -- |
| Configural v scalar | 1.5 | 5 | -- | -- | -- | 0.911 | -0.005 | 0.001 | 0.003 |
| Endline cross-arm (n=505) |  |  |  |  |  |  |  |  |  |
| Configural | 70.8 | 52 | 0.038 | 0.995 | 0.994 | -- | -- | -- | -- |
| Scalar | 74.1 | 57 | 0.034 | 0.996 | 0.995 | -- | -- | -- | -- |
| Configural v scalar | 2.9 | 5 | -- | -- | -- | 0.714 | -0.004 | 0.001 | 0.001 |
| Cross-time (B/E) (n=505) |  |  |  |  |  |  |  |  |  |
| Configural | 163.7 | 129 | 0.023 | 0.993 | 0.992 | -- | -- | -- | -- |
| Scalar | 178.1 | 145 | 0.021 | 0.994 | 0.993 | -- | -- | -- | -- |
| Configural v scalar | 20.0 | 16 | -- | -- | -- | 0.220 | -0.002 | 0.001 | 0.001 |
| Cross-time treatment (B/E) (n=237) |  |  |  |  |  |  |  |  |  |
| Configural** | 155.8 | 129 | 0.030 | 0.992 | 0.991 | -- | -- | -- | -- |
| Scalar*** | 173.1 | 145 | 0.029 | 0.992 | 0.991 | -- | -- | -- | -- |
| Configural v scalar | 22.5 | 16 | -- | -- | -- | 0.127 | -0.001 | 0.000 | 0.000 |
| Cross-time control (B/E) (n=268) |  |  |  |  |  |  |  |  |  |
| Configural | 137.4 | 129 | 0.016 | 0.996 | 0.995 | -- | -- | -- | -- |
| Scalar | 151.4 | 145 | 0.013 | 0.997 | 0.997 | -- | -- | -- | -- |
| Configural v scalar | 14.9 | 16 | -- | -- | -- | 0.533 | -0.003 | 0.001 | 0.002 |
| **OMC** | | | | | | | | | |
| **Model** | **χ2** | **d.f** | **RMSEA** | **CFI** | **TLI** | **Δχ2 (diff test)** | **ΔRMSEA** | **ΔCFI** | **ΔTLI** |
| Baseline cross-arm (n=1459) |  |  |  |  |  |  |  |  |  |
| Configural | 72.4 | 52 | 0.023 | 0.996 | 0.995 | -- | -- | -- | -- |
| Scalar | 74.4 | 57 | 0.020 | 0.997 | 0.996 | -- | -- | -- | -- |
| Configural v scalar | 2.3 | 5 | -- | -- | -- | 0.809 | -0.003 | 0.001 | 0.001 |
| Endline cross-arm (n=1458) |  |  |  |  |  |  |  |  |  |
| Configural | 72.3 | 52 | 0.023 | 0.999 | 0.999 | -- | -- | -- | -- |
| Scalar | 76.8 | 57 | 0.022 | 0.999 | 0.999 | -- | -- | -- | -- |
| Configural v scalar | 4.1 | 5 | -- | -- | -- | 0.534 | -0.001 | 0.000 | 0.000 |
| Cross-time (n=1460) |  |  |  |  |  |  |  |  |  |
| Configural | 170.1 | 129 | 0.015 | 0.998 | 0.998 | -- | -- | -- | -- |
| Scalar | 265.2 | 145 | 0.024 | 0.994 | 0.994 | -- | -- | -- | -- |
| Configural v scalar | 113.5 | 16 | -- | -- | -- | 0.000 | 0.009 | -0.004 | -0.004 |
| Cross-time treatment (n=746) |  |  |  |  |  |  |  |  |  |
| Configural | 143.3 | 129 | 0.012 | 0.999 | 0.999 | -- | -- | -- | -- |
| Scalar | 204.3 | 145 | 0.023 | 0.996 | 0.996 | -- | -- | -- | -- |
| Configural v scalar | 75.9 | 16 | -- | -- | -- | 0.000 | 0.011 | -0.003 | -0.003 |
| Cross-time control (n=714) |  |  |  |  |  |  |  |  |  |
| Configural | 138.0 | 129 | 0.010 | 0.999 | 0.999 | -- | -- | -- | -- |
| Scalar | 171.8 | 145 | 0.016 | 0.997 | 0.996 | -- | -- | -- | -- |
| Configural v scalar | 43.9 | 16 | -- | -- | -- | 0.000 | 0.006 | -0.002 | -0.003 |

**Online Supplement Table 7. Post Hoc Measurement Invariance Testing, Cross-time Treatment Condition, IND.**

| **IND (One-factor Model)** | | | | | | | | | |
| --- | --- | --- | --- | --- | --- | --- | --- | --- | --- |
| Model | χ2 | d.f. | RMSEA | CFI | TLI | Δχ2 (diff test) | ΔRMSEA | ΔCFI | ΔTLI |
| Cross-time treatment (B/E) (n=763) |  |  |  |  |  |  |  |  |  |
| Configural | 192.966 | 103 | 0.034 | 0.939 | 0.928 | -- | -- | -- | -- |
| Scalar | 245.941 | 119 | 0.037 | 0.913 | 0.913 | -- | -- | -- | -- |
| Configural v scalar | 73.608 | 16 |  |  |  | 0.0000 | 0.003 | -0.026 | -0.015 |
| Cross-time treatment (B/E) (n=763) Slap threshold freed |  |  |  |  |  |  |  |  |  |
| Configural | 192.966 | 103 | 0.034 | 0.939 | 0.928 | -- | -- | -- | -- |
| Scalar | 234.388 | 118 | 0.036 | 0.921 | 0.912 | -- | -- | -- | -- |
| Configural v scalar | 60.222 | 15 |  |  |  | 0.0000 | 0.002 | -0.018 | -0.016 |
| Cross-time treatment (B/E) (n=763) Force and Coerce correlated at baseline and endline |  |  |  |  |  |  |  |  |  |
| Configural | 121.727 | 101 | 0.016 | 0.986 | 0.983 | -- | -- | -- | -- |
| Scalar | 182.367 | 117 | 0.027 | 0.955 | 0.954 | -- | -- | -- | -- |
| Configural v scalar | 68.569 | 16 |  |  |  | 0.0000 | 0.011 | -0.031 | -0.029 |
| **IND (One-factor Model Without Coerce)** | | | | | | | | | |
| Model | χ2 | d.f. | RMSEA | CFI | TLI | Δχ2 (diff test) | ΔRMSEA | ΔCFI | ΔTLI |
| Cross-time treatment (B/E) (n=763) |  |  |  |  |  |  |  |  |  |
| Configural | 91 | 76 | 0.016 | 0.987 | 0.985 | -- | -- | -- | -- |
| Scalar | 146.416 | 89 | 0.029 | 0.952 | 0.951 | -- | -- | -- | -- |
| Configural v scalar | 61.171 | 13 |  |  |  | 0.0000 | 0.013 | -0.035 | -0.034 |
| Cross-time treatment (B/E) (n=763) Slap threshold freed |  |  |  |  |  |  |  |  |  |
| Model | χ2 | d.f. | RMSEA | CFI | TLI | Δχ2 (diff test) | ΔRMSEA | ΔCFI | ΔTLI |
| Configural | 91 | 76 | 0.016 | 0.987 | 0.985 | -- | -- | -- | -- |
| Scalar | 133.146 | 88 | 0.026 | 0.962 | 0.961 | -- | -- | -- | -- |
| Configural v scalar | 47.416 | 12 |  |  |  | 0.0000 | 0.01 | -0.025 | -0.024 |
| Cross-time treatment (B/E) (n=763) Slap threshold freed and slap loading freed |  |  |  |  |  |  |  |  |  |
| Model | χ2 | d.f. | RMSEA | CFI | TLI | Δχ2 (diff test) | ΔRMSEA | ΔCFI | ΔTLI |
| Configural | 91 | 76 | 0.016 | 0.987 | 0.985 | -- | -- | -- | -- |
| Scalar | 127.72 | 87 | 0.025 | 0.966 | 0.964 | -- | -- | -- | -- |
| Configural v scalar | 41.475 | 11 |  |  |  | 0.0000 | 0.009 | -0.021 | -0.021 |
| **IND (One-factor Model Without Othsex)** | | | | | | | | | |
| Cross-time treatment (B/E) (n=763) |  |  |  |  |  |  |  |  |  |
| Model | χ2 | d.f. | RMSEA | CFI | TLI | Δχ2 (diff test) | ΔRMSEA | ΔCFI | ΔTLI |
| Configural | 173.918 | 76 | 0.041 | 0.938 | 0.926 | -- | -- | -- | -- |
| Scalar | 227.776 | 90 | 0.045 | 0.913 | 0.912 | -- | -- | -- | -- |
| Configural v scalar | 78.063 | 14 |  |  |  | 0.0000 | 0.004 | -0.025 | -0.014 |
| Cross-time treatment (B/E) (n=763) Coerce and force correlated at endline |  |  |  |  |  |  |  |  |  |
| Model | χ2 | d.f. | RMSEA | CFI | TLI | Δχ2 (diff test) | ΔRMSEA | ΔCFI | ΔTLI |
| Configural | 145.536 | 75 | 0.035 | 0.956 | 0.946 | -- | -- | -- | -- |
| Scalar | 201.434 | 89 | 0.041 | 0.929 | 0.928 | -- | -- | -- | -- |
| Configural v scalar | 75.532 | 14 |  |  |  | 0.0000 | 0.006 | -0.027 | -0.018 |

**Online Supplement Table 8. Post Hoc Measurement Invariance Testing, OMC.**

| **OMC (One-factor Model)** | | | | | | | | | |
| --- | --- | --- | --- | --- | --- | --- | --- | --- | --- |
| Model | χ2 | d.f | RMSEA | CFI | TLI | Δχ2 (diff test) | ΔRMSEA | ΔCFI | ΔTLI |
| Baseline cross-arm (n=1459) |  |  |  |  |  |  |  |  |  |
| Configural | 173.389 | 54 | 0.055 | 0.979 | 0.972 | -- | -- | -- | -- |
| Scalar | 177.777 | 61 | 0.051 | 0.98 | 0.976 | -- | -- | -- | -- |
| Configural v scalar | 8.256 | 7 | -- | -- | -- | 0.3106 | -0.004 | 0.001 | 0.004 |
| Endline cross-arm (n=1458) |  |  |  |  |  |  |  |  |  |
| Configural | 214.417 | 54 | 0.064 | 0.995 | 0.993 | -- | -- | -- | -- |
| Scalar | 220.702 | 61 | 0.06 | 0.995 | 0.994 | -- | -- | -- | -- |
| Configural v scalar | 7.927 | 7 | -- | -- | -- | 0.3391 | -0.004 | 0 | 0.001 |
| Cross-time (n=1460) |  |  |  |  |  |  |  |  |  |
| Configural | 335.563 | 134 | 0.032 | 0.99 | 0.989 | -- | -- | -- | -- |
| Scalar | 409.133 | 151 | 0.034 | 0.988 | 0.987 | -- | -- | -- | -- |
| Configural v scalar | 122.988 | 17 | -- | -- | -- | 0.0000 | 0.002 | -0.002 | -0.002 |
| Cross-time treatment (n=746) |  |  |  |  |  |  |  |  |  |
| Configural | 215.926 | 134 | 0.029 | 0.994 | 0.993 | -- | -- | -- | -- |
| Scalar | 266.148 | 151 | 0.032 | 0.992 | 0.992 | -- | -- | -- | -- |
| Configural v scalar | 82.261 | 17 | -- | -- | -- | 0.0000 | 0.003 | -0.002 | -0.001 |
| Cross-time control (n=714) |  |  |  |  |  |  |  |  |  |
| Configural | 204.316 | 134 | 0.027 | 0.991 | 0.99 | -- | -- | -- | -- |
| Scalar | 233.586 | 151 | 0.028 | 0.989 | 0.989 | -- | -- | -- | -- |
| Configural v scalar | 49.332 | 17 | -- | -- | -- | 0.0001 | 0.001 | -0.002 | -0.001 |
